# Protection by vaccines and previous infection against the Omicron variant of SARS-CoV-2

**DOI:** 10.1101/2022.02.24.22271396

**Authors:** Martin Šmíd, Luděk Berec, Ondřej Májek, Tomáš Pavlík, Jiří Jarkovský, Jakub Weiner, Lenka Přibylová, Tamara Barusová, Jan Trnka

## Abstract

The Omicron variant of the SARS-CoV-2 virus carries mutations, which enable it to evade immunity conferred by vaccines and previous infections. We used a Cox proportional hazards model and a logistic regression model on individual-level data on all laboratory-confirmed SARS-CoV-2 infections in the Czech Republic to estimate the relative risk of infection, hospitalization, including severe states, for Delta and Omicron variants, adjusting for sex, age, previous infection, vaccine type and vaccination status. A recent (<2 months) two-dose vaccination reached VE 43% (95% CI: 42-44) against infection by Omicron compared to 73% (95% CI: 72-74) against Delta. A recent booster increased VE to 56% (95% CI: 55-56) against Omicron infection compared to 90% (95% CI: 90-91) for Delta. The VE against Omicron hospitalization of a recent two-dose vaccination was 45% (95% CI: 29-57), with a recent booster 87% (95% CI: 84-88). The VE against the need for oxygen therapy due to Omicron was 57% (95% CI: 32-72) for recent vaccination, 90% (95% CI: 87-92) for a recent booster. Post-infection protection against Omicron hospitalization declined from 68% (95% CI: 68-69) at <6 months to 13% (95% CI: 11-14) at >6 months after a previous infection. A recent combination of a previous infection and vaccination was more protective then either alone with a slight benefit from a vaccination preceding an infection. Once infected, the OR for Omicron relative to Delta was 0.36 (95% CI: 0.34-0.38) for hospitalization, 0.24 (95% CI: 0.22-0.26) for oxygen therapy, and 0.24 (95% CI: 0.21-0.28) for ICU admission.

**Significance Statement:** A nation-wide study shows that the protection of a previous infection or vaccination is lower against Omicron compared to Delta variant of SARS-CoV-2 and further declines with time. A booster dose or a combination of post-infection immunity with a vaccine conferred significant benefit to individuals in the Omicron wave in the Czech Republic, which further strengthens the importance of vaccination as an effective public health measure.

---

The B.1.1.529 (Omicron) variant of SARS-CoV-2 was first detected in South Africa in November 2021, immediately designated a variant of concern by the WHO (1) and thereafter seen quickly to spread throughout most of the world. This rapid spread was at least in part brought about by a degree of immune evasion due to a large number of mutations in the viral S-protein, which led to changes in epitopes recognised by antibodies elicited by vaccination or previous infection (2). Together with non-pharmacological interventions such as face masks, distancing, ventilation of interior spaces testing and isolating, vaccination is among the most effective means of individual and collective protection from the impacts of the pandemic. The immune evasion by the Omicron variant thus caused concern and led to a lot of interest in both laboratory and real-life epidemiological data that could accurately measure this phenomenon.

Since December 27, 2020 the inhabitants of the Czech Republic have been receiving covid-19 vaccines with the largest number vaccinated with the mRNA vaccine Comirnaty (BNT162b2, Pfizer/BioNTech), followed by Spikevax (mRNA-1273, Moderna), and the adenovirus-based vector vaccines Vaxzevria (ChAdOx1 nCoV-19, AstraZeneca) and Janssen Covid-19 Vaccine (Ad26.CoV2.S, Johnson&Johnson, further abbreviated as Janssen) (3). By February 13, 2022, the end of our study period, 68% of the population had a complete vaccination and 39% received a booster dose (3)

The first case of the Omicron variant in the Czech Republic was detected at the end of November 2021, its proportion of recorded cases rapidly rose and by January 10, 2022 it became the dominant variant, see Figure 1. An increasing number of infections among fully vaccinated and re-infections indeed suggests that immune evasion poses a significant risk to further covid-19 development (3).

**Fig. 1.**
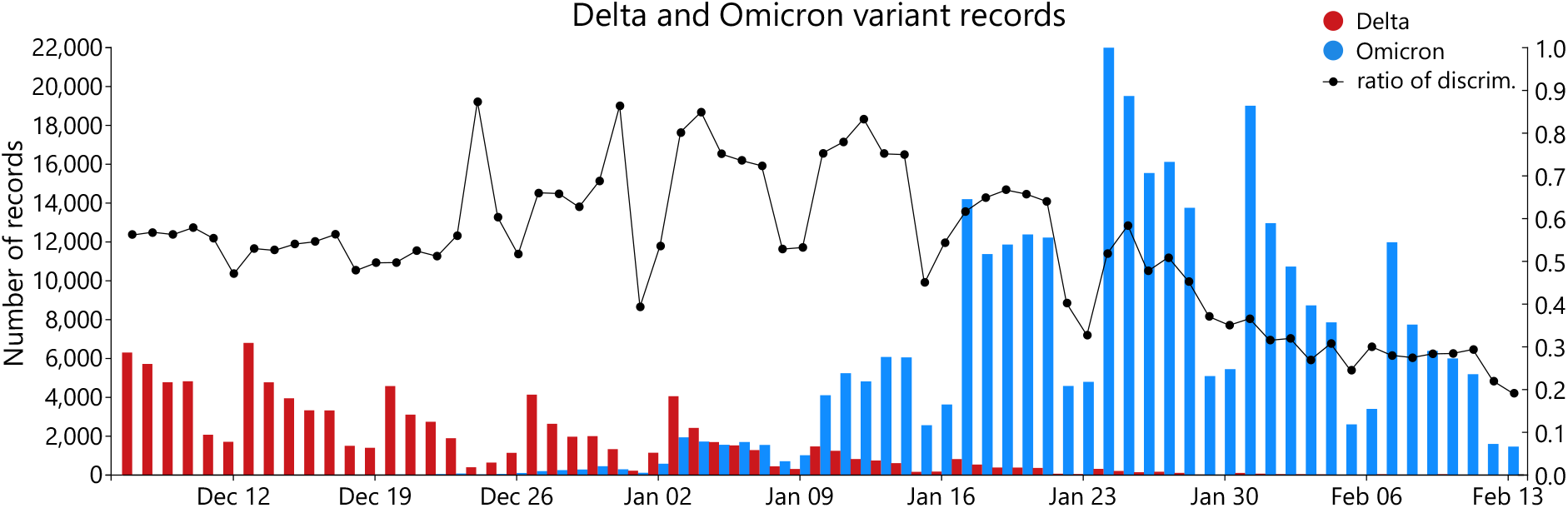
Number of recorded cases with assigned Delta (red) and Omicron (blue) variant and the proportion of PCR positive tests (black) tested for viral variants using multiplex PCR.

In this study, we estimate how the protection due to vaccination or previous SARS-CoV-2 infection against covid-19 infection, hospital admission, oxygen therapy and ICU admission varies in relation to the virus variant and time elapsed for the whole population of the Czech Republic.

## Methods

### Study population and data sources

The analyses are based on data from the Czech National Information System of Infectious Diseases (ISID), which includes records of all individuals tested positive for SARS-CoV-2 in the Czech Republic since the beginning of covid-19 pandemic, including children (4). This database is overseen by the Czech Ministry of Health and operated by the Institute of Health Information and Statistics of the Czech Republic. Data are routinely collected in compliance with Czech legal regulations (Act on the Protection of Public Health). The Director of the Institute of Health Information and Statistics of the Czech Republic has granted an ethical approval of the retrospective analyses presented in this paper.

The ISID database collects demographic data (age, sex and region of residence), dates of vaccination, including the vaccine types for each dose, and dates of infection and potential reinfection, hospitalization including treatment type and the date of potential death with covid-19. Approximately half of data recorded in the study period include information on whether the infection is caused by the Omicron, Delta, some other variant, or that a variant discrimination was not performed. The information on the Omicron variant is based on results of multiplex PCR or viral genome sequencing, which are available only for a subset of all PCR-positive cases. The identification of Delta, Omicron, or other variant was computed using definition of viral S-protein mutations according to ECDC (5); the algorithm was tailored to multiplex PCR kits used in the Czech Republic in collaboration with the National Institute of Public Health and the National Reference Laboratory (6). Additional information on deaths from any cause come from the Death Certificate System; these data are used for censoring purposes only.

### Study endpoints

We studied four types of events: (i) SARS-CoV-2 infection defined as a PCR-confirmed positive test of any type of sample regardless of the presence of symptoms, (ii) hospital admission of a person, who tested positive on a PCR test, within two weeks after the confirmed infection or earlier, (iii) use of any type of oxygen therapy (nasal oxygen, noninvasive ventilation, invasive mechanical ventilation, high-flow nasal oxygen, and extracorporeal membrane oxygenation) and (iv) admission to ICU during the hospitalization. All events were related to the date of infection report.

We examined events during the two month period from December 7, 2021 to February 13, 2022 during which Delta and Omicron switched dominance in the Czech Republic (Fig. 1).

### Statistical analysis

A Cox regression with time-varying covariates was applied to estimate hazard ratios (HRs) for the outcomes of interest separately for each viral variant. In these analyses the infections by the variant other than the examined one and the infections lacking variant assignment were censored at the time of infection. Analogously to (7) we used calendar time instead of time from event occurrence as the time scale. Thus the time course of individual cases was modelled by means of “switching” dummy variables, corresponding to the development of the immune status after vaccination or past infection in 61-day periods for vaccination and 121-day periods for the time from the last infection.

The protection provided by vaccine (vaccine effectiveness) or previous infection is calculated by comparing hazards of the vaccinated and/or immunized individuals to those of the “control group” – those who have not been vaccinated and infected so far and subtracted from 1 using the equation:

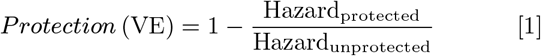

Further we examine the post-infection immunity by estimating hazard ratios of infection of previously unvaccinated individual in relation to time elapsed from the infection. By using calendar time we were able automatically to incorporate the changing conditions of the epidemic, including non-pharmacological measures, seasonal effects, and the virus variant, as all of these phenomena can be included in the underlying baseline hazard function.

To examine the probabilities of hospitalization, oxygen therapy and ICU admission for an infected individual we use the logistic regression with the event of interest as the outcome and with all infections with an assigned viral variant as the data. By means of the dummy corresponding to the virus variant we compare the probabilities of the outcome for both the variants. Post-infection and post-vaccination status, age and sex are used as control variables. To estimate the amount of protection provided by the vaccination and/or past infection we run the logistic regression separately for infections by the two variants.

All calculations were performed using the R software (package `survival`). The algorithm used to transform data from the database into the package command inputs was coded in C++. See Supplementary material 1 for details.

## Results

### Protection against infection

First we looked at the protection conferred by vaccination or a previous infection against a new infection, since the protection against infection represents the potential to protect others at risk in the population. For vaccination and the Omicron variant the protection reached 43% (95% CI 42-44) shortly after completing the full vaccination scheme, falling to 9% (95% CI 8-10) after more than two months. This protection increased to 56% (95% CI 55-56) shortly after receiving a booster dose, followed by a decline to 21% (95% CI 19-23) after more than two months. These numbers strongly contrast with the protection against the Delta variant, which was consistently higher at 73% (95% CI 72-74), 57% (95% CI 56-58), 90% (95% CI 90-91) and 82% (95% CI 79-84), respectively. Similar degrees of protection against infection are conferred also by the post-infection immunity: 68% (95% CI 68-69) shortly after infection (2-6 months, a positive test during the first two months after an infection is not considered a reinfection by definition) and 13% (95% CI 11-14) after six months for Omicron, versus 95% (95% CI 94-96) shortly after infection and 83% (95% CI 82-84) after six months for Delta (Figure 2). Based on the past prevalence of viral variants it can be expected that the infections older than 6 months were mostly due to the original Wuhan, D614G and Alpha variants, while the more recent ones were predominantly due to Delta. As we show in the Supplementary material 2, Sections 11 and 12, explicit accounting for the vaccine type Comirnaty (BNT162b2, Pfizer/BioNTech) and Spikevax (mRNA-201273, Moderna) gave values of effectiveness comparable with the analyses of pooled data reported here in the main text.

**Fig. 2.**
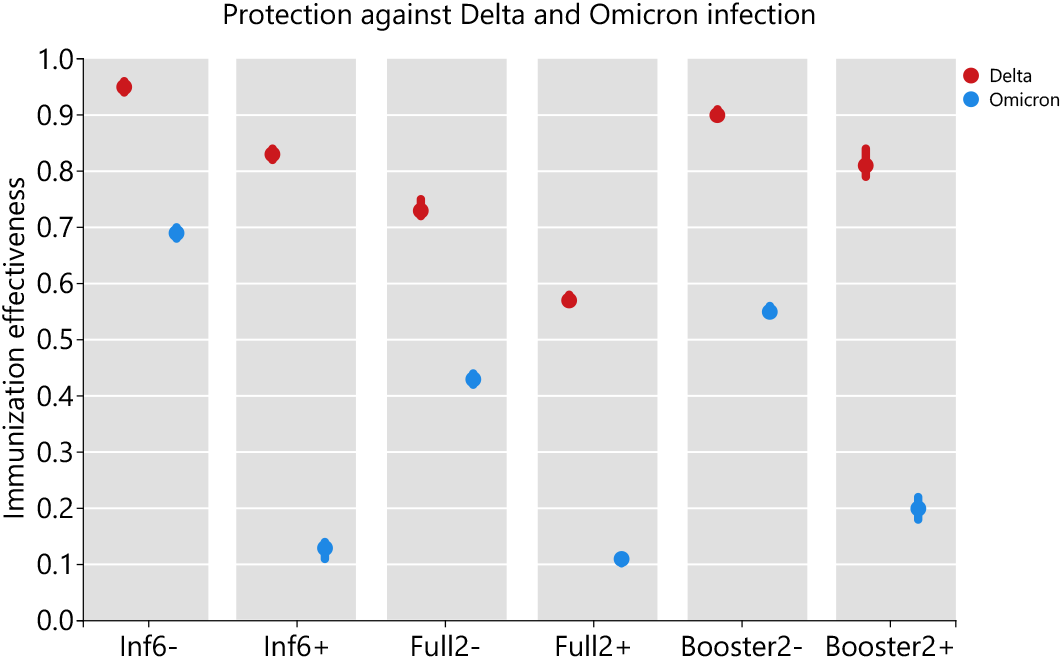
Protection provided by vaccination or previous infection against infection by the Omicron and Delta variants of the SARS-CoV-2 virus. Inf6-, previous infection <6 months ago; Inf6+, previous infection >6 months ago; Full2-, complete vaccination <2 months ago; Full2+, complete vaccination >2 months ago; Booster2-, booster dose <2 months ago; Booster2+, booster dose >2 months ago. Shown are point estimates of protection with 95% CI.

We had enough data to examine all the combinations in which a previous infection preceded vaccination. As expected, protection declined with time elapsed from the previous infection or vaccination (Tables 1 and 2). For the Delta variant, any combination provided ≥ 95% protection against infection (Table 2). This protection remained quite high also for Omicron when the previous infection was recent, falling to lower values for an older previous infection, but even then the protection was significantly higher than for a vaccination or previous infection alone (Table 1). We also analysed cases, where a vaccination preceded a previous infection, and conclude that when the protection is high (against Delta) then the order of events does not matter (see description of Tab. 2). However, when protection is lower (against Omicron), the situation when a previous infection followed a vaccination appeared to provide a higher protection than the inverse sequence (see the caption of Tab. 1). The protection provided by Full 2+/Inf 6-combination was 89% (88-91) as compared to 86% (95% CI 85-88) for Inf 6-/Full 2+.

**Table 1.**
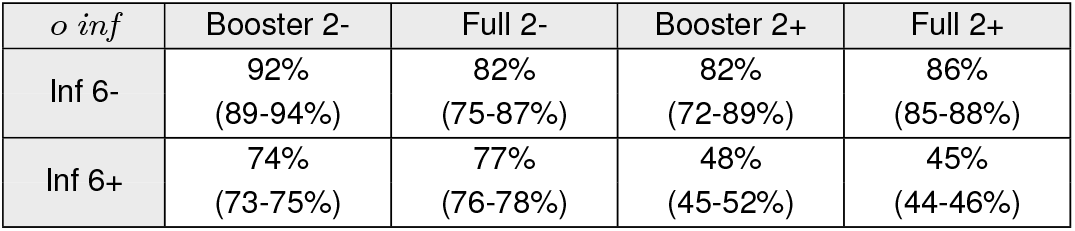
Protection due to various combinations of past infection preceding vaccination *against infection* for the *Omicron* variant of the SARS-CoV-2 virus. Infection in recent 6 months is denoted as Inf 6- and older Inf 6+, completed vaccination scheme is denoted Full 2- for completion in recent 2 months and Full 2+ for older completion, analogously for booster. In parentheses, 95% confidence intervals (CI) are given. The inverse immunisation order had a higher effect: more than 2 months old full vaccination followed by infection in recent 6 months had 89% (88-91%) for full vaccination.

**Table 2.**
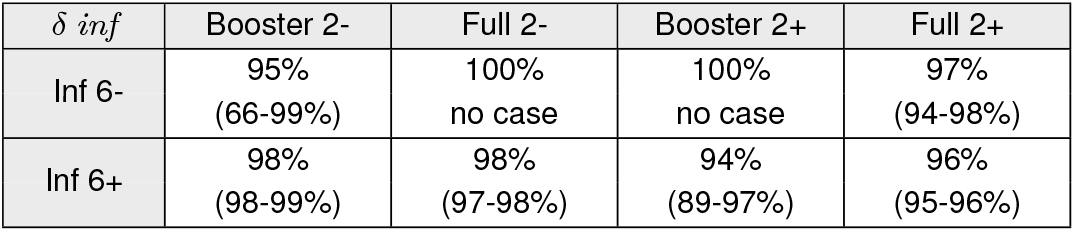
Protection due to various combinations of past infection preceding vaccination *against infection* for the *Delta* variant of the SARS-CoV-2 virus, 95% confidence intervals (CI) in parentheses. The inverse immunisation order: more than 2 months old full vaccination followed by infection in recent 6 months had protection 96% (90-98%) for full vaccination.

| $\delta inf$ | Booster 2- | Full 2- | Booster 2+ | Full 2+ |
| --- | --- | --- | --- | --- |
| Inf 6- | 95%<br>(66-99%) | 100%<br>no case | 100%<br>no case | 97%<br>(94-98%) |
| Inf 6+ | 98%<br>(98-99%) | 98%<br>(97-98%) | 94%<br>(89-97%) | 96%<br>(95-96%) |

These levels of protection against infection provided by vaccination or previous infection were estimated from the complete set of data. A finer grained analysis of temporal dynamics of immunity waning after a previous infection was then conducted specifically for individuals that were previously infected but remained non-vaccinated. For Omicron, the protection was estimated as 69% (95% CI 68-69) for 2-6 months after previous infection, 48% (95% CI 46-50) for 7-10 months, 34% (95% CI 33-35) for 11-14 months, and 17% (95% CI 15-18) for 14 and more months after previous infection. For Delta, on the contrary, these numbers were 93% (95% CI 91-94), 91% (95% CI 90-92), 86% (95% CI 85-86), and 79% (95% CI 77-81), respectively.

### Protection against hospitalization

Qualitatively similar pattern yet quantitatively consistently higher protection is seen against hospitalization (Tables 3), a need for oxygen therapy (Tables 6), and a need for intensive care (Tables 7). For example, a recent booster dose provides 87% protection against hospitalization, 90% against a need of oxygen therapy, and 84% against a need of intensive care for the Omicron variant. Moreover, all combinations of previous infection and vaccination present in our data appear to provide nearly complete protection against Omicron as regards hospitalization (Tables 4 and 5) as well as against need of oxygen therapy or intensive care (no cases have been observed for such situations).

**Table 3.** Vaccine effectiveness and protection provided by post-infection immunity *against hospitalization*, for the Omicron and Delta variants of the SARS-CoV-2 virus, 95% confidence intervals (CI) in parentheses.

| Effect ag. Hosp. | Omicron | Delta |
| --- | --- | --- |
| Full 2- | 45% (29-57%) | 75% (68-80%) |
| Full 2+ | 29% (21-37%) | 79% (78-81%) |
| Booster 2- | 87% (84-88%) | 98% (97-98%) |
| Booster 2+ | 79% (75-83%) | 97% (95-98%) |
| Inf 6- | 87% (73-94%) | 100% (no case) |
| Inf 6+ | 92% (86-96%) | 95% (93-96%) |

**Table 4.**
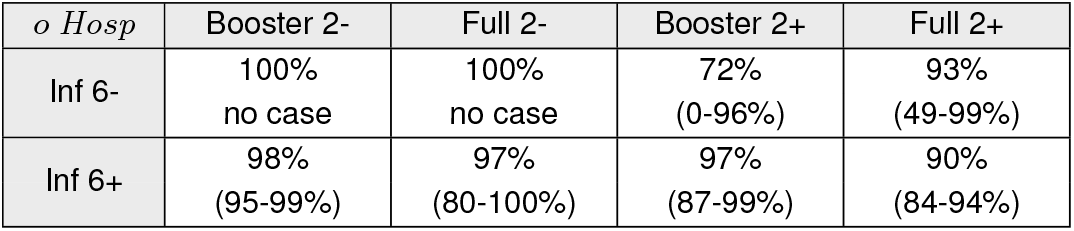
Protection due to various combinations of past infection preceding vaccination *against hospitalization* for the *Omicron* variant of the SARS-CoV-2 virus, 95% confidence intervals (CI) in parentheses. The inverse immunisation order: more than 2 months old full vaccination followed by infection in recent 6 months had protection 94% (78-99%)

**Table 5.** Protection due to various combinations of past infection preceding vaccination *against hospitalization* for the *Delta* variant of the SARS-CoV-2 virus, 95% confidence intervals (CI) in parentheses. The inverse immunisation order: more than 2 months old vaccination followed by infection in recent 6 months had protection 100% (no case) for full vaccination.

| *δ Hosp* | Booster 2- | Full 2- | Booster 2+ | Full 2+ |
| --- | --- | --- | --- | --- |
| Inf 6- | 100%<br>no case | 100%<br>no case | 100%<br>no case | 100%<br>no case |
| Inf 6+ | 100%<br>(99-100%) | 98%<br>(95-99%) | 98%<br>(86-100%) | 98%<br>(98-99%) |

**Table 6.** Vaccine effectiveness and protection provided by post-infection immunity against hospitalization with a *need for oxygen therapy*, for the Omicron and Delta variants of the SARS-CoV-2 virus, 95% confidence intervals (CI) in parentheses.

| Effect ag. $O_2$ | Omicron | Delta |
| --- | --- | --- |
| Full 2- | 57% (32-72%) | 82% (76-87%) |
| Full 2+ | 32% (20-43%) | 82% (80-83%) |
| Booster 2- | 90% (87-92%) | 98% (98-98%) |
| Booster 2+ | 85% (80-88%) | 97% (95-98%) |
| Inf 6- | 81% (40-94%) | 100% (no case) |
| Inf 6+ | 88% (72-94%) | 98% (95-99%) |

**Table 7.** Vaccine effectiveness and protection provided by post-infection immunity against hospitalization with a need for *intensive care*, for the Omicron and Delta variants of the SARS-CoV-2 virus, 95% confidence intervals (CI) in parentheses.

| Effect ag. ICU | Omicron | Delta |
| --- | --- | --- |
| Full 2- | 58% (3-82%) | 84% (72-91%) |
| Full 2+ | 37% (12-55%) | 86% (83-88%) |
| Booster 2- | 83% (75-89%) | 98% (97-99%) |
| Booster 2+ | 60% (37-74%) | 97% (92-99%) |
| Inf 6- | 83% (0-98%) | 100% (no case) |
| Inf 6+ | 66% (15-86%) | 97% (90-99%) |

### Risk of a severe outcome for Omicron vs. Delta

Finally, our logistic regression analyses show that once infected, the odds ratio for hospitalization with Omicron relative to Delta is 0.36 (0.34-0.38), for a need of oxygen therapy with Omicron relative to Delta is 0.24 (0.22-0.26), and for a need of intensive care with Omicron relative to Delta is also 0.24 (0.21-0.28). Moreover, once hospitalized, the odds ratio for a need of oxygen therapy with Omicron relative to Delta is 0.44 (0.39-0.49) and for a need of intensive care with Omicron relative to Delta is 0.64 (0.52-0.72).

## Discussion and Conclusions

Our data support the existing evidence that the Omicron variant of SARS-CoV-2 evades to a significant extent both the post-vaccination and post-infection immunity (2, 8–11). The VEs of all the vaccines used in the Czech Republic are lower for Omicron compared to Delta. As we previously observed with Alpha and Delta (12), the protection against infection by the Omicron variant wanes over time too. However, a booster vaccine dose provides robust and lasting or slowly waning protection against hospitalization, the need for oxygen therapy and intensive care. The combined post-infection and post-vaccination immunity is the most protective regardless of the exact sequence of events, suggesting that the best protective strategy before a coming wave is to vaccinate all individuals, whether previously vaccinated or with a previous covid-19 infection.

We are aware of the complicated interpretation of the hospitalization data for the Omicron wave, as the very high basic reproduction number *R*_0_ of this variant (13) translated into the very high prevalence of infection in the population at the peak of the epidemic wave and a much higher proportion of hospitalized patients with covid-19 as a concomitant finding rather than the reason for admission. We therefore analysed separately the need for oxygen therapy and ICU admission as a more relevant measure of severe outcomes due to the Omicron infection.

Compared to the Delta variant, the protection provided by the post-infection or post-vaccination immunity is lower for the Omicron variant but at the same time the Omicron variant appears less severe than the Delta variant and the odds ratio for oxygen therapy or ICU admission are both approximately about one quarter compared to the Delta variant.

A common limitation of studies like ours is the fact that only a certain portion of infections is reported (ascertainment rate). We believe this phenomenon does not affect significantly our estimates of vaccine effectiveness, assuming that the ascertainment rate is the same for the vaccinated and the unvaccinated alike and we have no evidence to the contrary. A difference in the ascertainment rate could also distort our estimates of the protection by the post-infection immunity; in particular, if there were undetected individuals with post-infection immunity in the control group, the infection risk of the population would be underestimated and, consequently, the protection by infection underestimated as well. Our results should be interpreted in terms of reported infections only.

## Data Availability

Data are not publicly available but may be (partially) obtained from Institute of Health Information and Statistics of the Czech Republic for research purposes.

## ACKNOWLEDGMENTS

Data reported in this study and used for the analyses are not public. De-identified individual-level data are available to the scientific community. Requests should be submitted to the Institute of Health Information and Statistics of the Czech Republic (www.uzis.cz/index-en.php), together with a short description of their analysis proposals, where they will be assessed based on relevance and scientific merit.

## Supplementary material 1: Protection provided by covid-19 vaccines and previous infection against the Omicron variant

### Dataset

In total, our dataset contains 8,282,080 records of vaccinated and/or SARS-CoV-2 positive persons, out of which 10,937 were excluded for either inconsistency or the lack of valid sex and/or age information. Additional 97,855 records were excluded as corresponding individuals died before 7. December 2021 which we set as the start date (34,145 were recorded to die from covid-19, 63,740 from other reasons; see Table S3). As the source dataset consists only of those who were tested positive and/or were vaccinated, we added subjects who were neither tested positive nor vaccinated. In particular, we completed each sex-age category to the numbers reported by the Czech Statistical Office by December 31, 2020 – 10,701,777 inhabitants; consequently, our sample truly reflected the sex and age structure of the whole population, containing all the positive and/or vaccinated individuals. We neglected births and deaths of the added persons. Moreover, not all non-covid deaths were recorded as they are reported with a delay.

### Methods: a Cox regression with time-varying covariates

For the analyses of vaccine effectiveness and immunity protection, made separately for both the Omicron and Delta variant, we use the Cox proportional hazards model with time-varying covariates.

The input of the model consists of one or more records for each subject. Each record corresponds to a time interval from time `T1` to `T2`, in which the covariates are constant and in the interior of which none of the outcomes happened. Each record contains indicators of outcomes `VariantInf, VariantHosp, VariantOxygen, VariantICU, DeadByCov, DeadByOther`, happening at *T* 2 and the value of the covariates, valid in [*T* 1, *T* 2). Undiscriminated infections and infections by other variants lead to withdrawal from the study (right censoring). Deaths of covid (`DeadByCov`) or of other reasons (`DeadByOthe`r) lead to right censoring at the time of the event as well. Fixed (non-time-dependent) covariates include sex (`Sex`) and age category (`AgeGr`). Time is measured in days and we take the day before our study period (Dec 7th, 2021) as time zero. By default, for each subject, the intervals cover the entire period from 7th. December 2021 to 13. February 2022. The period is shortened (censoring takes place) if either:

- The subject is infected (except for the Infection analysis)
- The subject is infected by some other variant or the infection is not discriminated (in the Infection analysis)
- The subject is reported to die
- The subject is reported to obtain booster by Astra or Janssen
- The subject gets a vaccine (only in the reinfection analysis)

#### Combined Immunity

In these analyses, we let each individual to go through the several “immunity states” encoded into `Immunity` covariate. The outcomes (events) are either (a confirmed) infection (`VariantInf`, may be repeated), hospitalization (`VariantHosp`), oxygen therapy (`VariantOxygen`) or ICU admission (`VariantICU`).

The `Immunity` categorical covariate can take the following values:

`_noimmunity` meaning that the subject was neither vaccinated nor underwent (comfirmed) infection,

*X*_alone meaning that the condition *X* happened,

*X*_*Y* which means that the condition *X* happened, being followed by condition *Y* in time.

The conditions X and Y could be:

`part` partially vaccinated (from 14 days of the dose application)

**Table S1.** Details on analyses by Cox regression.

| Analysis | Outcome | Covariates |
| --- | --- | --- |
| Infections | VariantInf | Immunity, Sex, AgeGr |
| Reinfections | VariantInf | InfPrior, Sex, AgeGr |
| Hospitalizations | VariantHosp | Immunity, Sex, AgeGr |
| Oxygen therapy | VariantOxygen | Immunity, Sex, AgeGr |
| ICU admission | VariantICU | Immunity, Sex, AgeGr |
| Detailed hospitalizations | VariantHosp | VaccStatus, InfPrior, Sex, AgeGr |
| Detailed oxygen therapy | VariantOxygen | VaccStatus, InfPrior, Sex, AgeGr |
| Detailed ICU admission | VariantICU | VaccStatus, InfPrior, Sex, AgeGr |

`full2`-fully vaccinated no earlier than two months ago (from 14 days to 14 + 61 *−* 1 = 74 days after the dose application)

`full2+` fully vaccinated earlier than two months ago (from 75 days from the application or earlier)

`boost2-` booster vaccinated no earlier than two months ago (from 14 days to 74 days after the dose application)

`boost2+` booster vaccinated earlier than tow months ago (from 75 days from the application or earlier)

`inf6-` underwent infection no earlier than six months (180 days) ago

`inf6+` underwent infection earlier than six months (180 days) ago.

#### Re-infection Analysis

When separately estimating the post-infection immunity we consider the infection by the examined variant (`VariantInf`) as the outcome. As the explanatory variable we take the categorical covariate `InfPrior` indicating the delay from the last infection, grouped by four months. The `InfPrior` covariate can take the following values:

`_noinf` no previous infection

`inf1` 60–180 days after the last confirmed infection (the re-infection cannot happen during the first 60 days by definition)

`inf2` 181–301 days after the last confirmed infection

`inf3` 302–422 days after the last confirmed infection

`inf4+` any later.

#### Detailed Analyses

Here, the outcomes include `VariantInf` (possibly repeated), `VariantHosp` or `VariantOxygen`. The time-varying covariates include `VaccStatus` and `InfPrior`. The `VaccStatus` is either `_novacc` (not vaccintaed) or takes the form *VSP* where

*V* denotes the vaccine (P-fizer, M-oderna, A-stra, J-anssen),

*S* denotes the state of vaccination (part - partial, full - full, boost - booster)

*P* denotes the time distance from the last vaccination

**S=part:** 1=first 61 days after the dose takes effect (14 days after application), 2=any later

**S=full:** 1=first 61 days after the dose takes effect (14 days after application), 2=next 61 days, 3=later

**S=boost:** 1=first 61 days after the dose takes effect (7 days after application), 2=later

The `InfPrior` covariate is as above.

Table S1 summarizes all the analyses we made by means of Cox regression.

### Methods: a Logistic regression

In theses analyses, the binary outcomes `Hosp, Oxygen, ICU` corresponding to the discriminated infections are explained by dummy `Variant` taking values either `Omicron` or `Delta`, the value of covariate `Immunity` valid at the time of the infection and constant covariates `AgeGr` anad `Sex`. Table S2 summarizes all these analyses.

### Software implementation

The `coxph` function from `survival R` package was used for the Cox regression, the (`glm` function from `stats R` package) for the Logistic regression. The original software package was used to transform the source data to the input of the functions. The source code with example (unreal) data can be retrieved from https://github.com/bisop-repo/omicronprotection.

### Supplementary tables

**Table S2.**
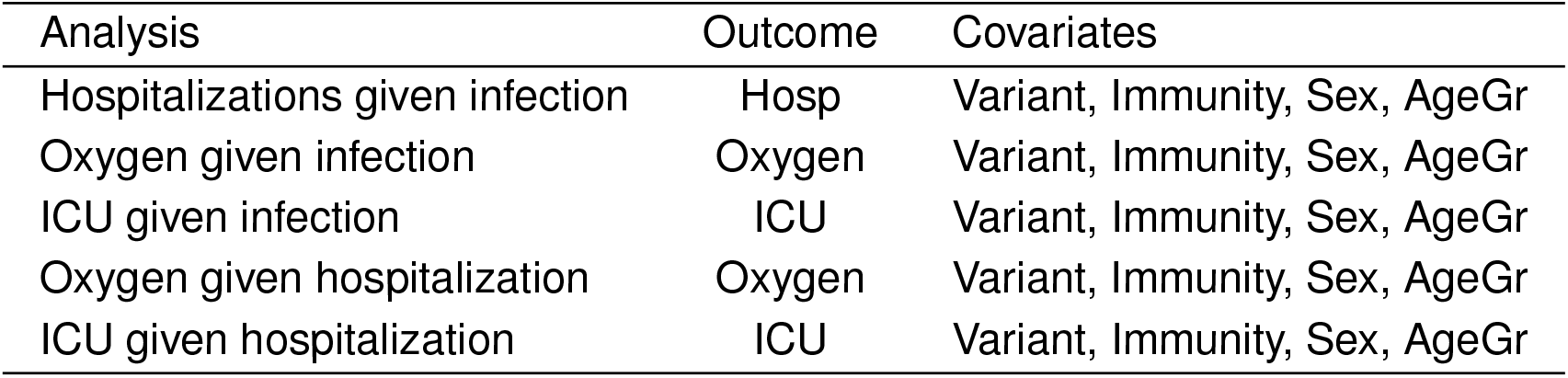
Details on analyses by Logistic regression.

**Table S3.**
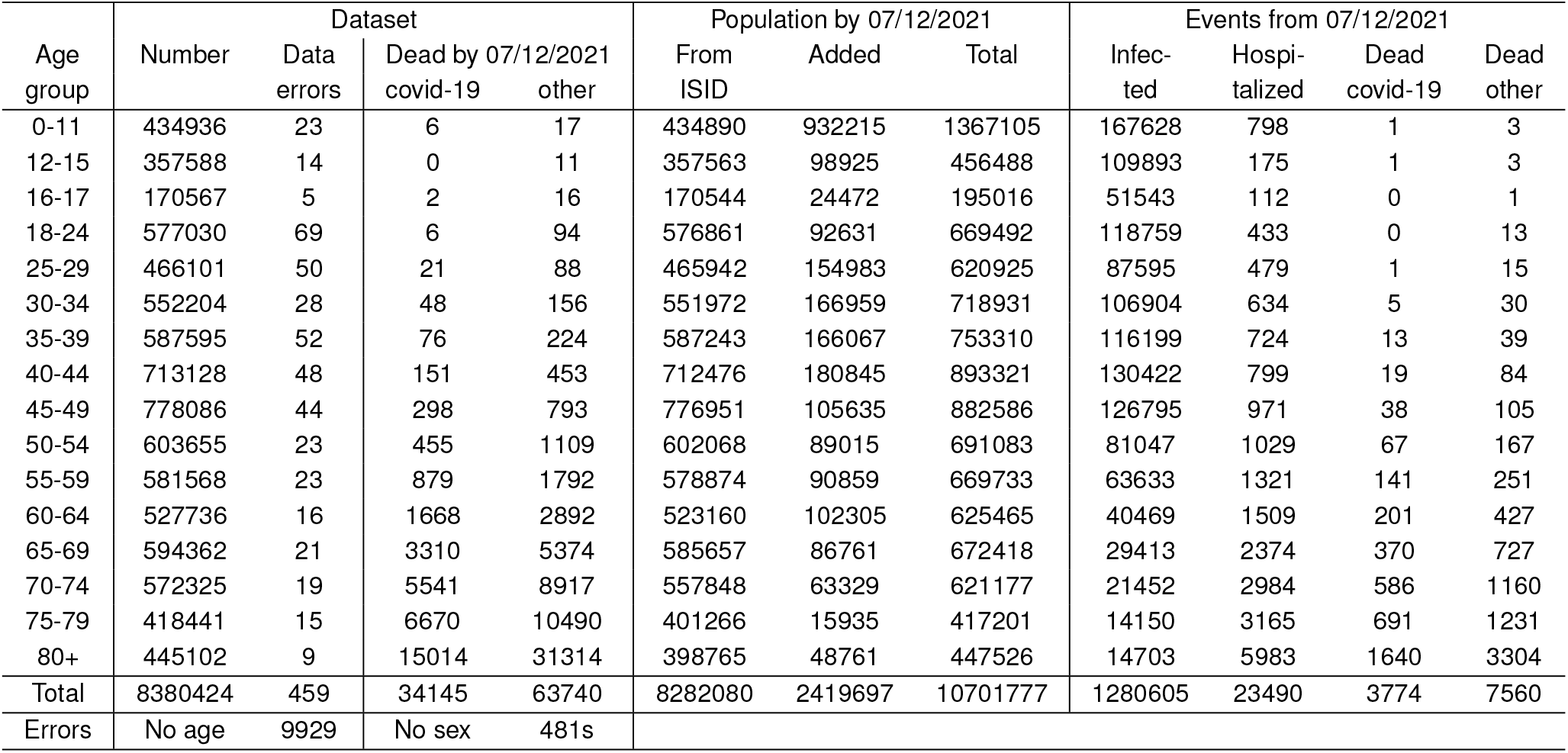
Descriptive statistics: population and epidemic characteristics in the Czech Republic; ISID = the Czech National Information System of Infectious Diseases.

**Table S4.**
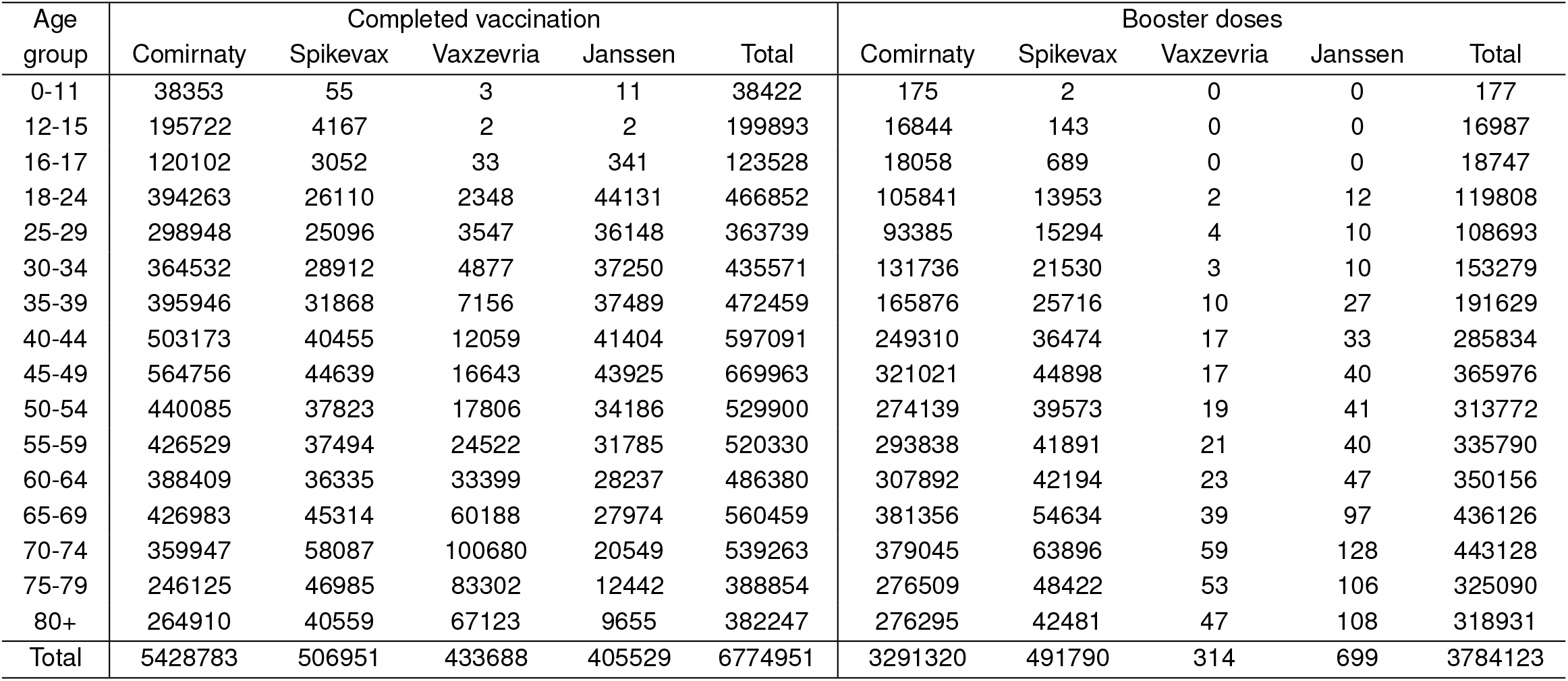
Descriptive statistics: vaccine distribution among different age groups until November 20, 2021.

## Results of Statistical Estimation

### 1 Infections – Omicron

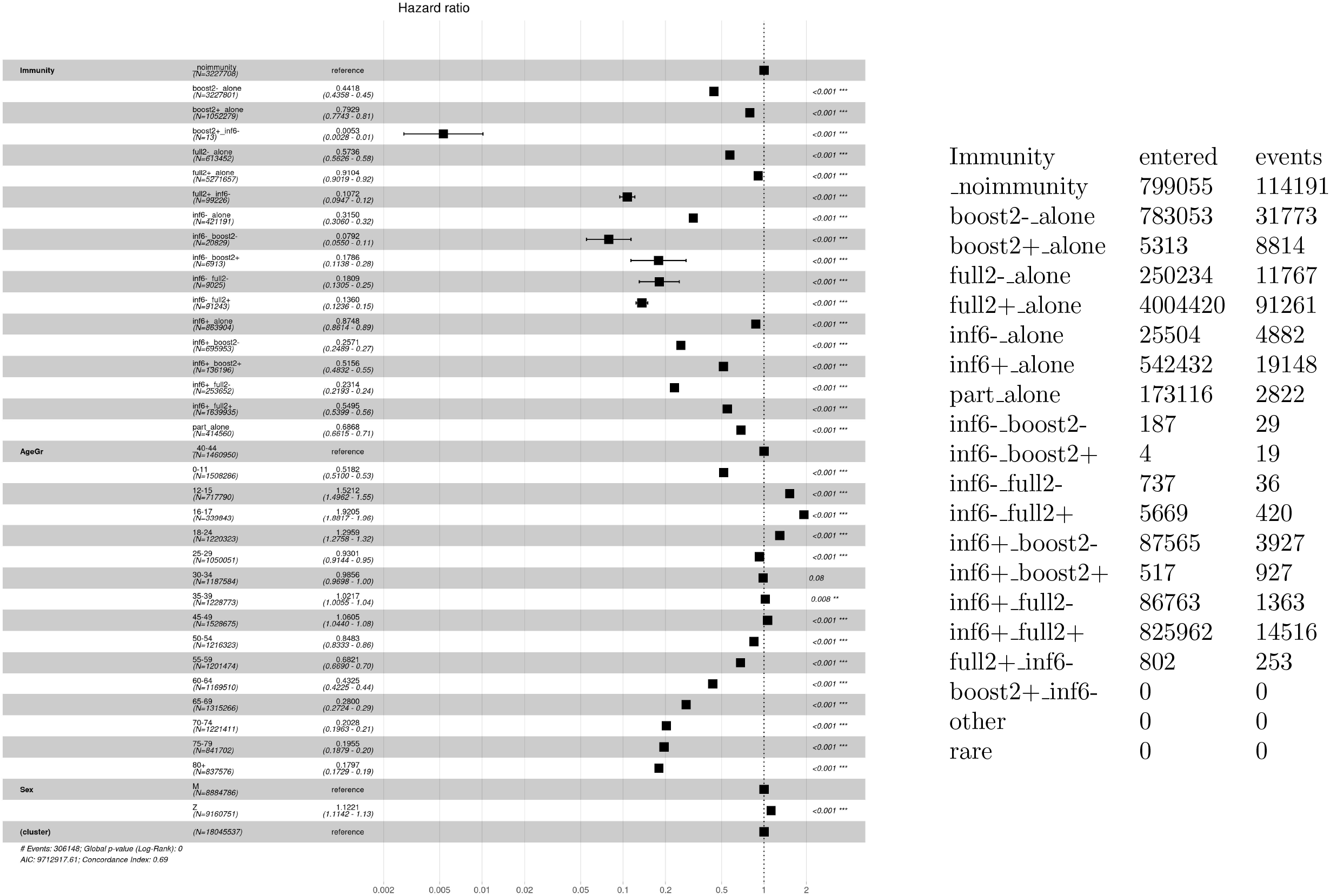

### 2 Infections – Delta

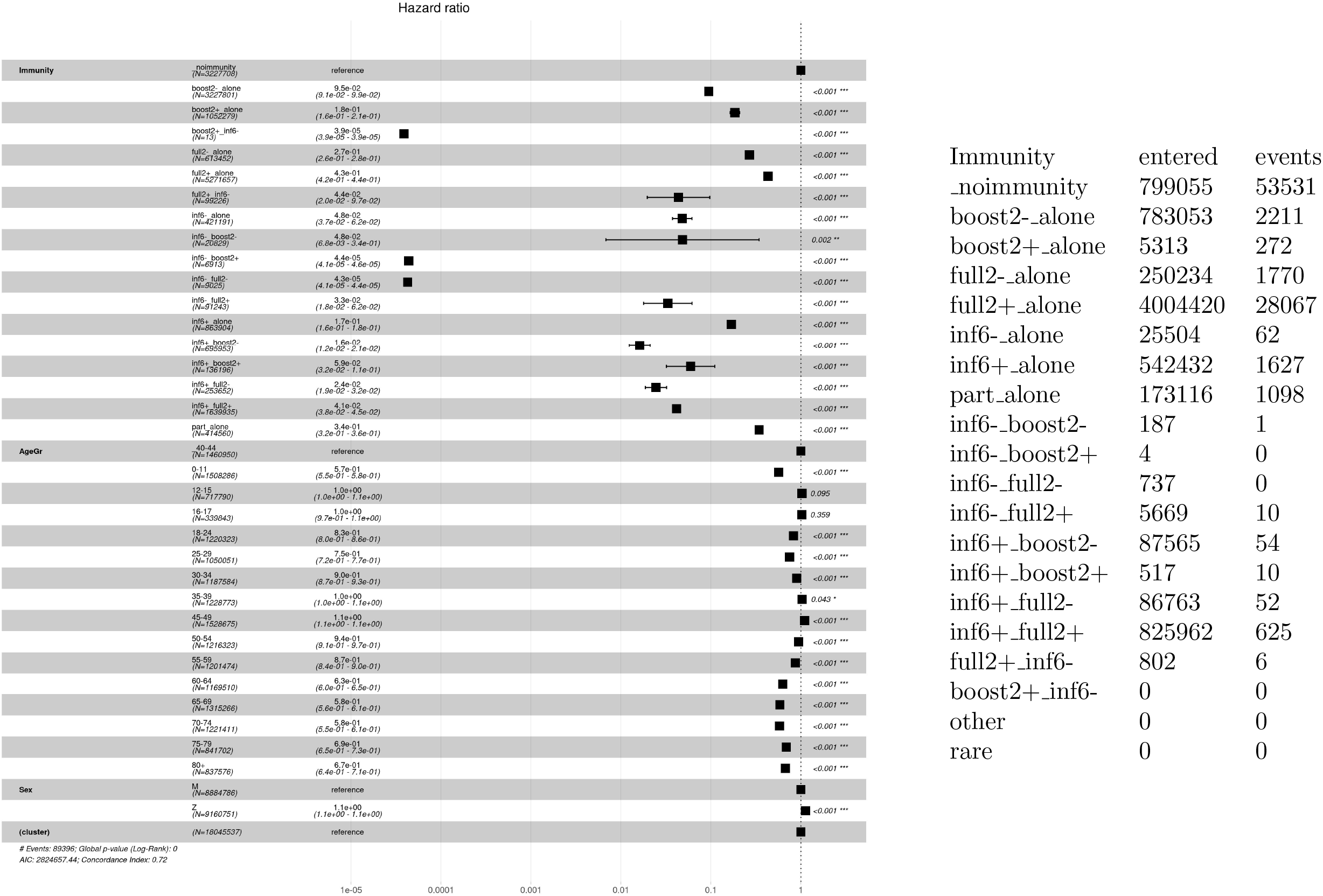

### 3 Re-infections – Omicron

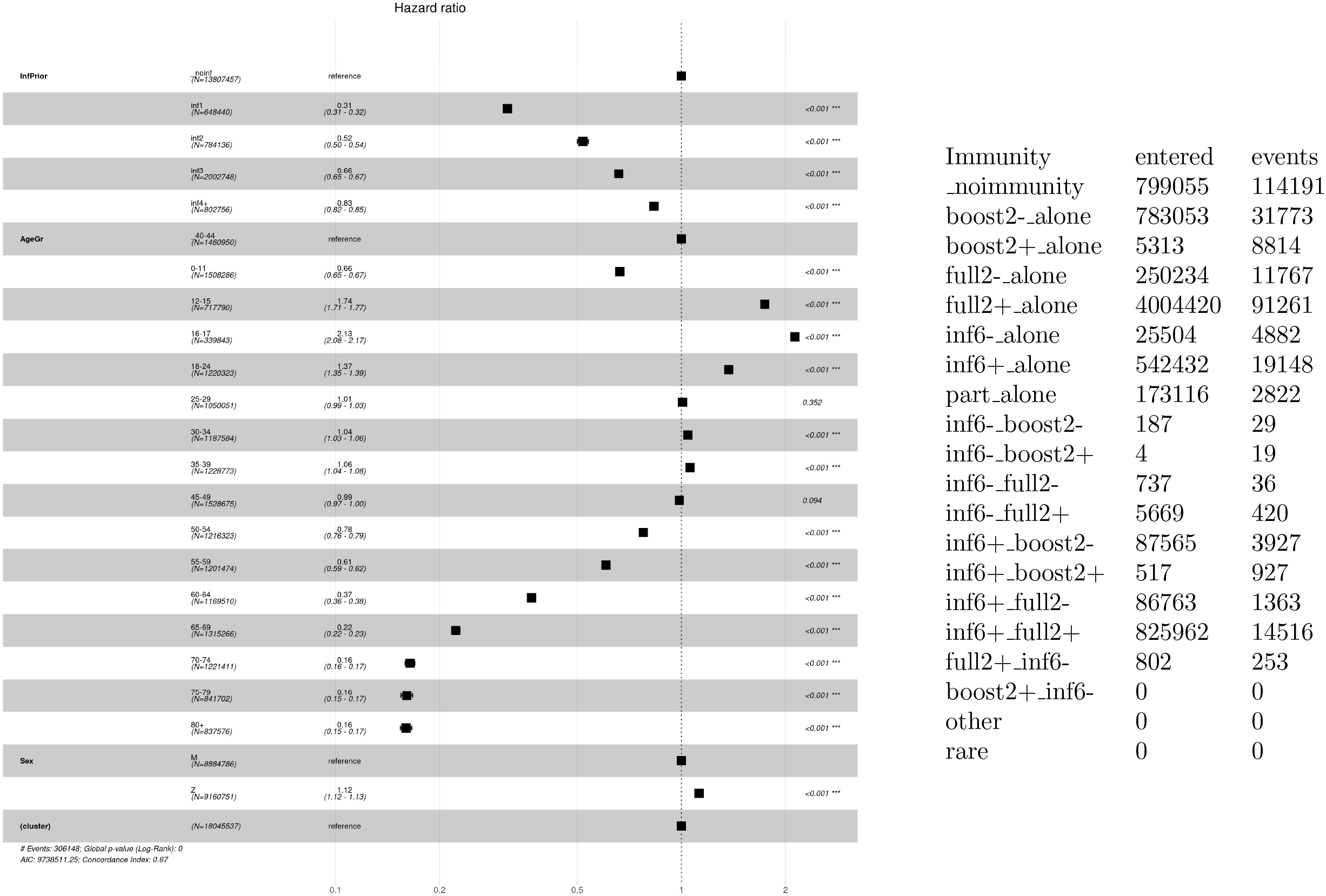

### 4 Re-infections – Delta

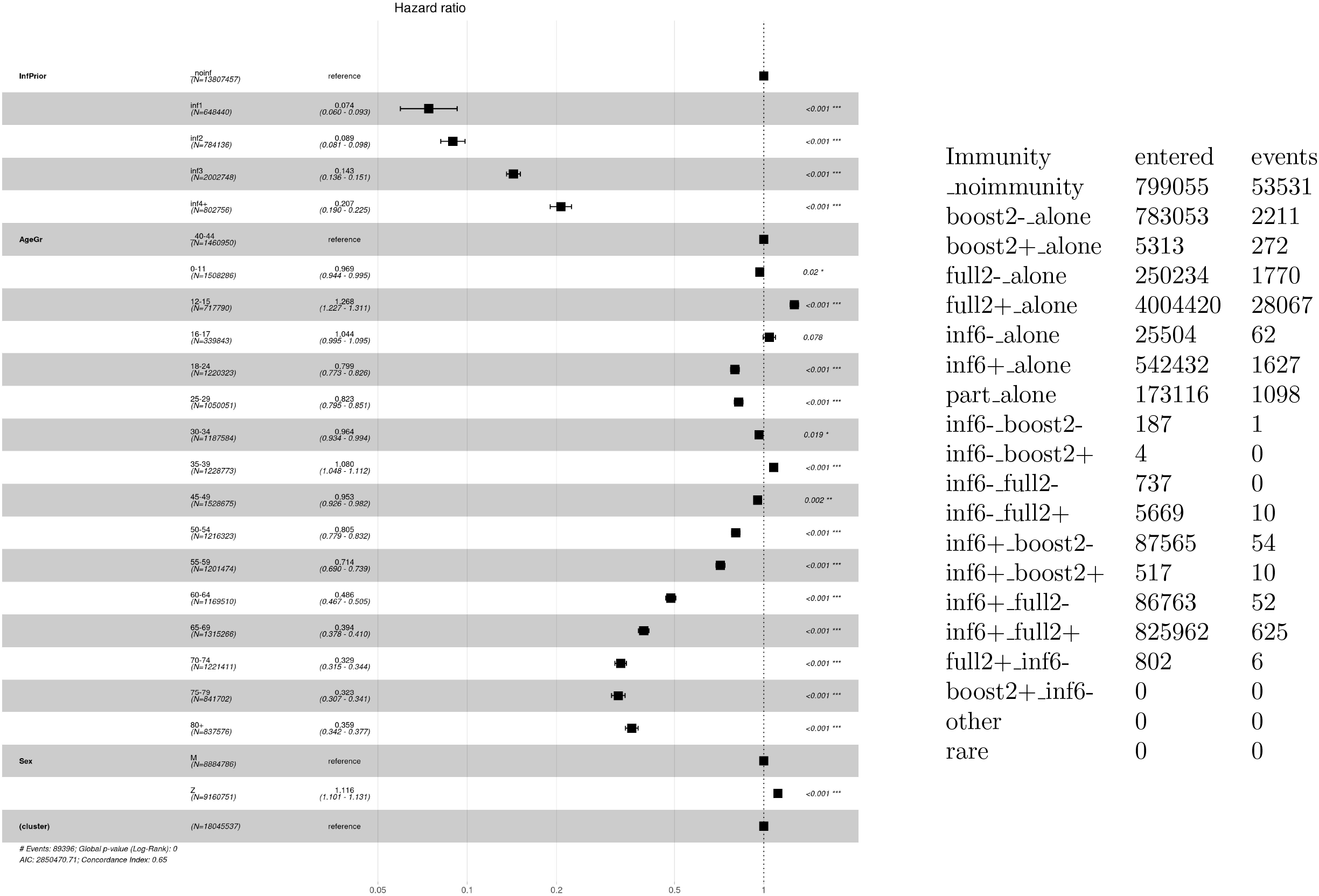

### 5 Hospitalizations – Omicron

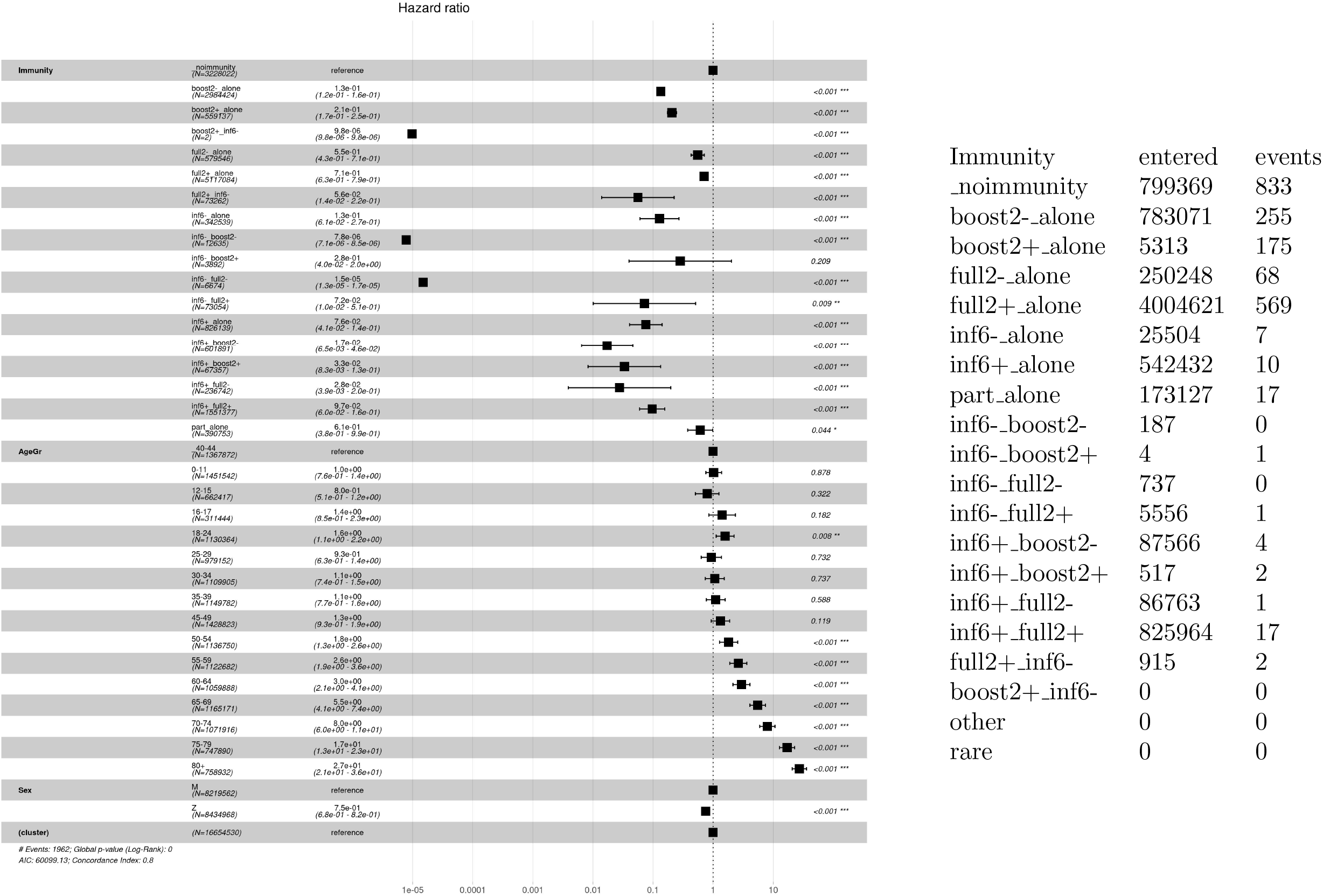

### 6 Hospitalizations – Delta

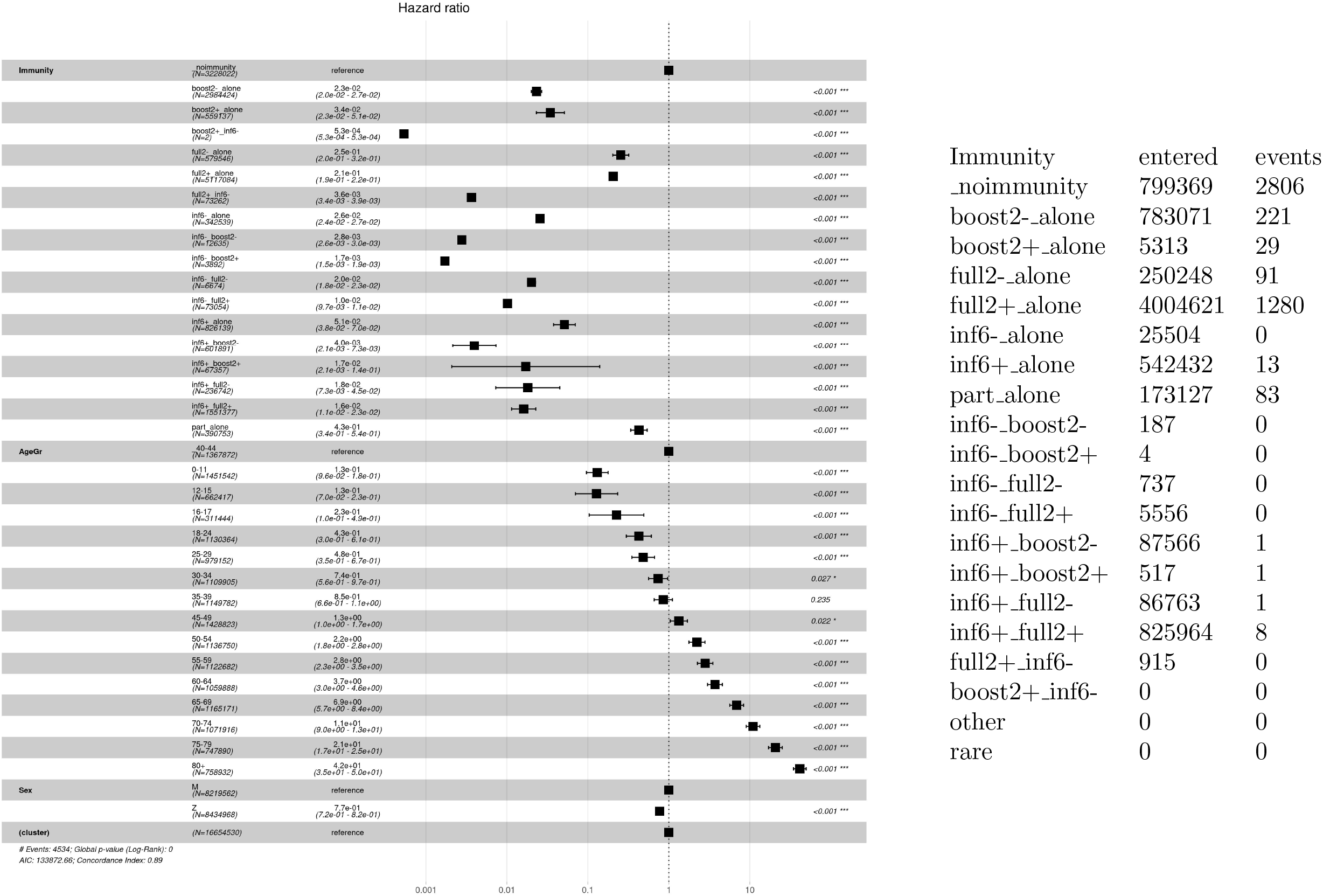

### 7 Oxygen therapy – Omicron

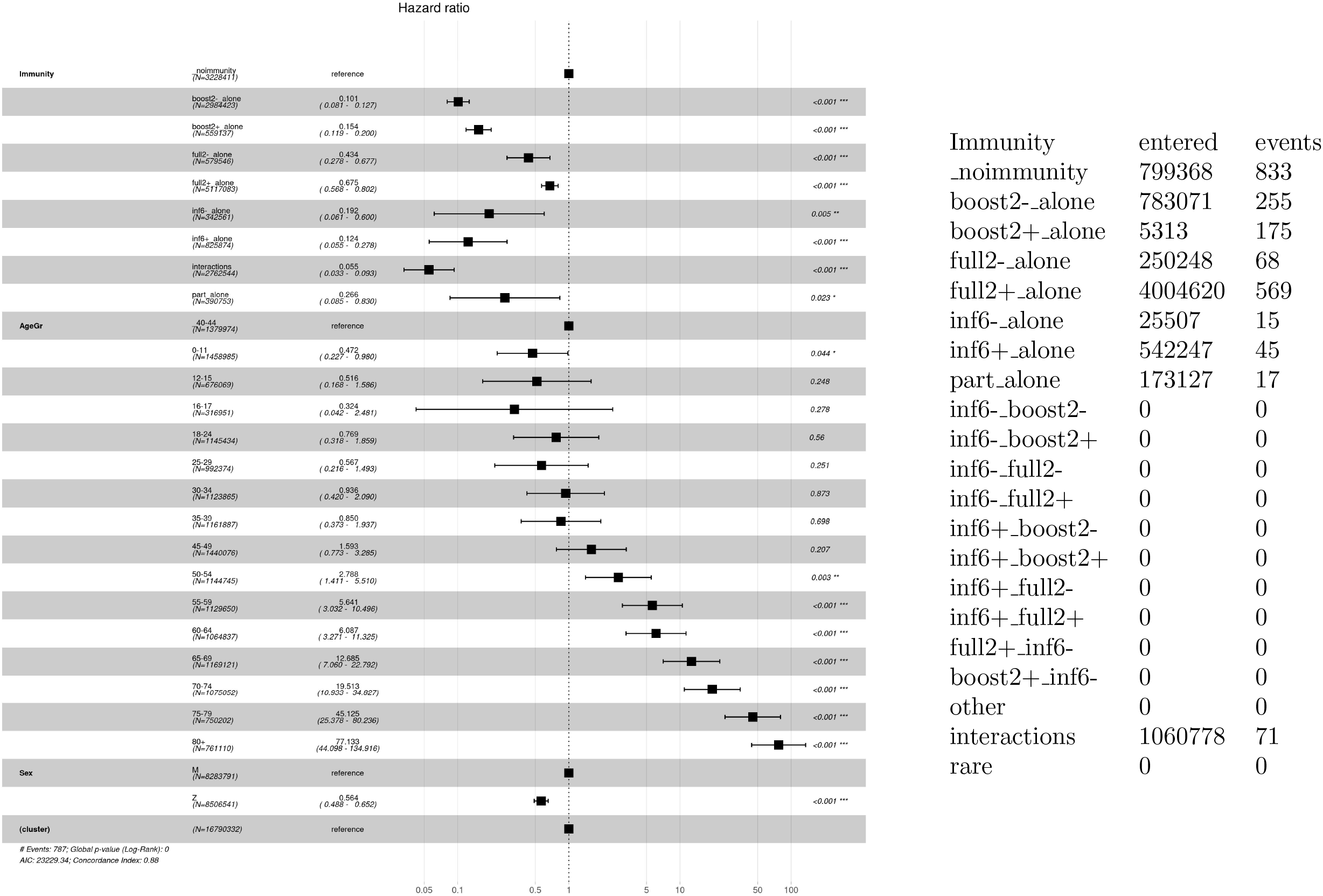

### 8 Oxygen therapy – Delta

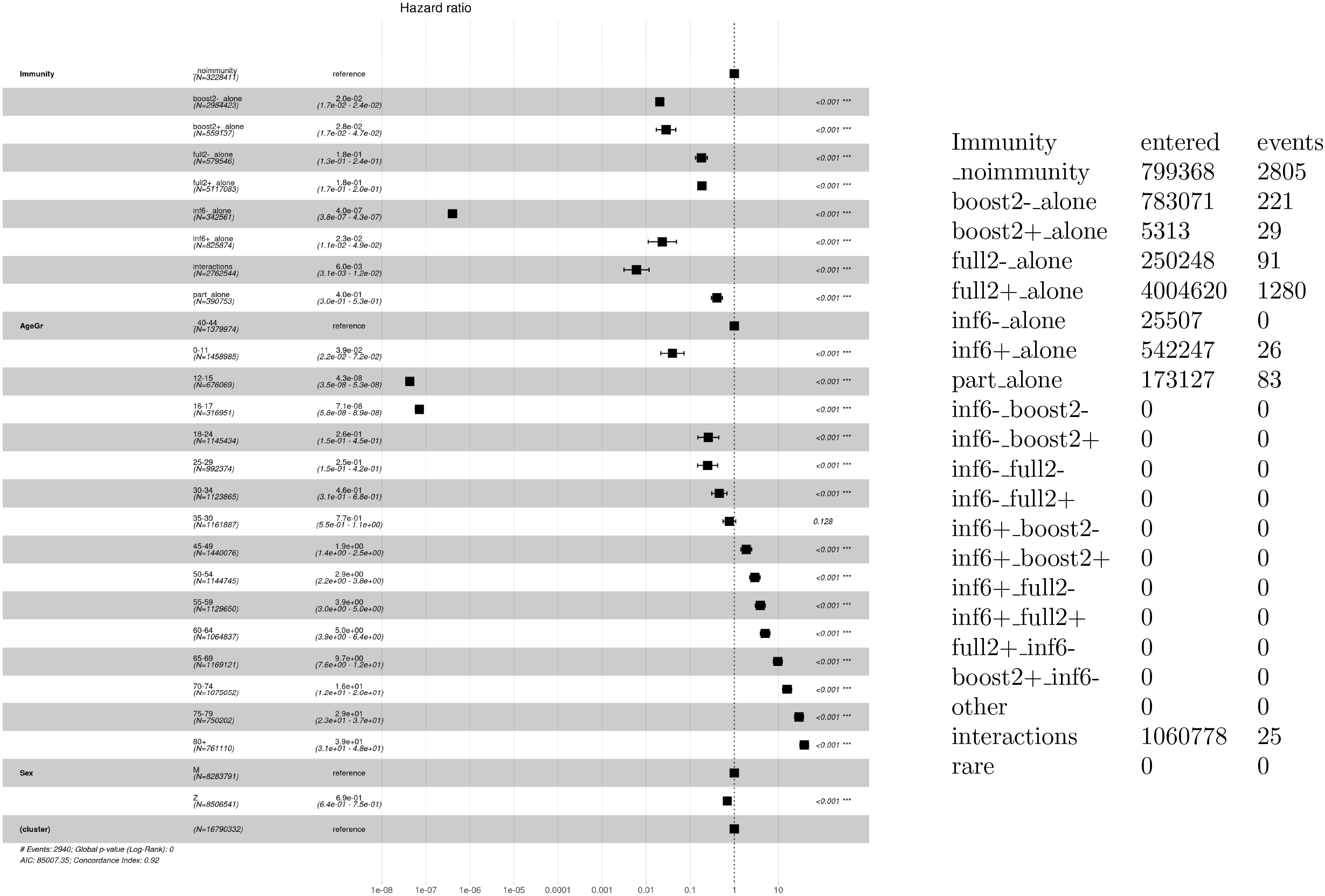

### 9 ICU therapy – Omicron

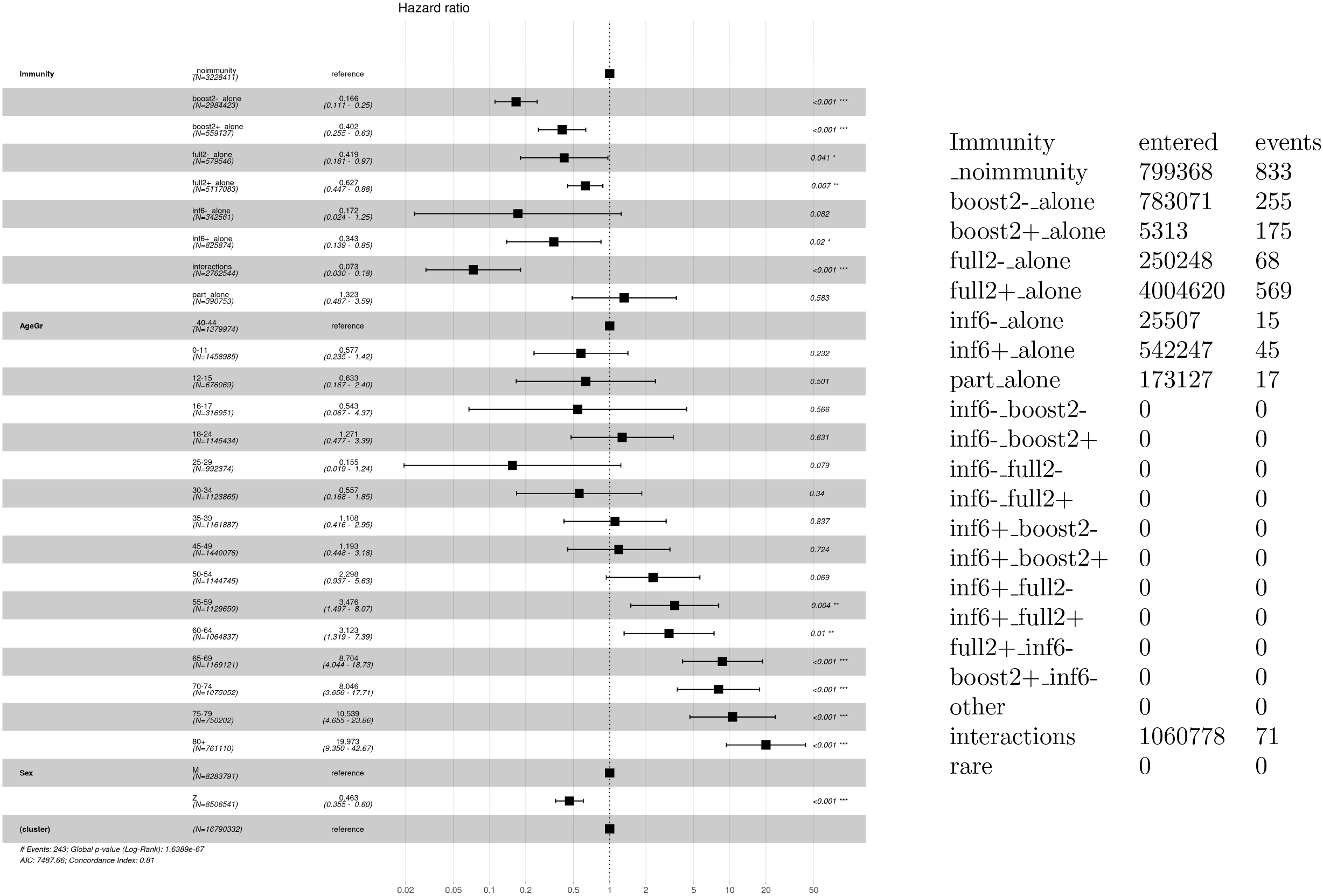

### 10 ICU therapy – Delta

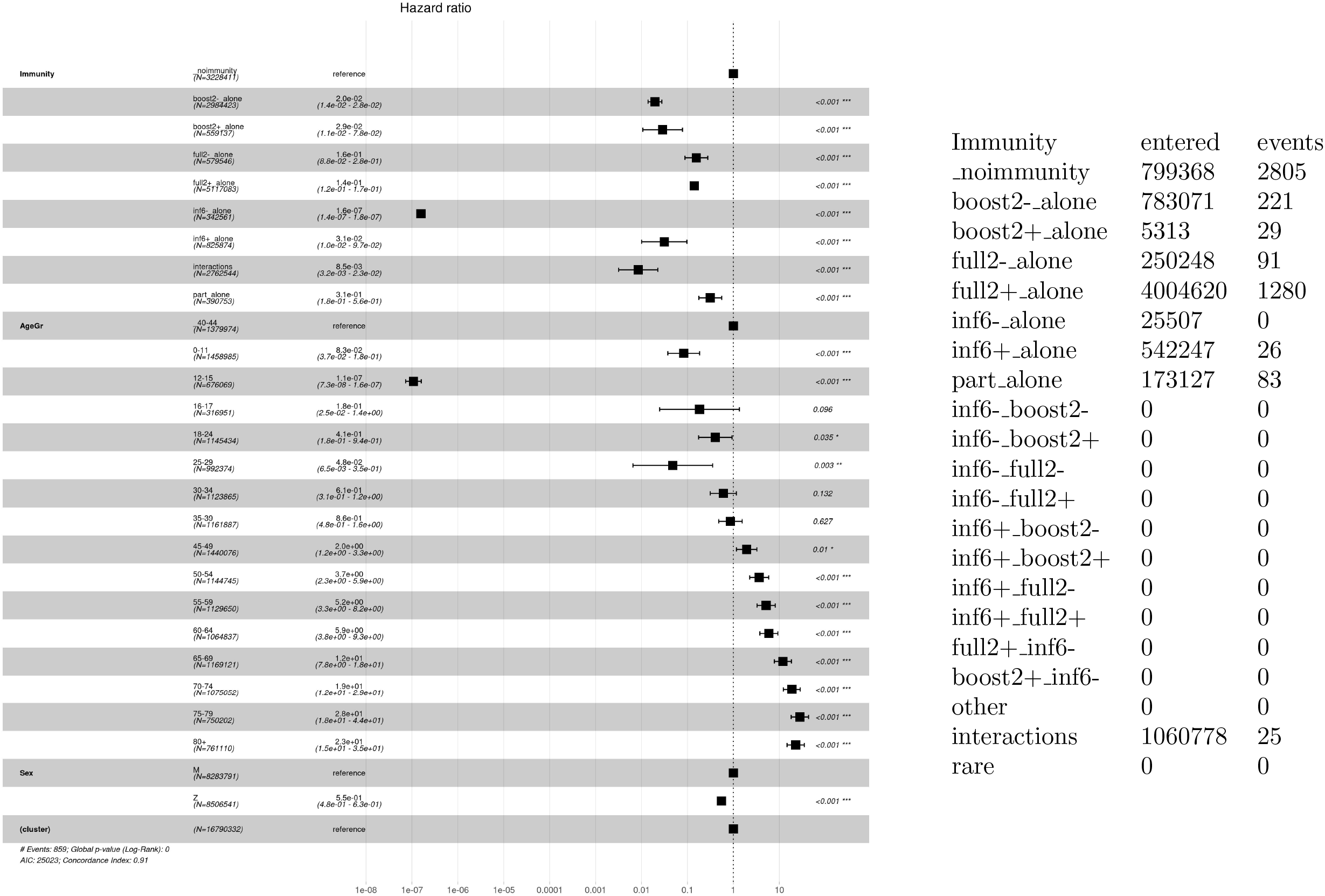

### 11 Infections by Vaccines - Omicron

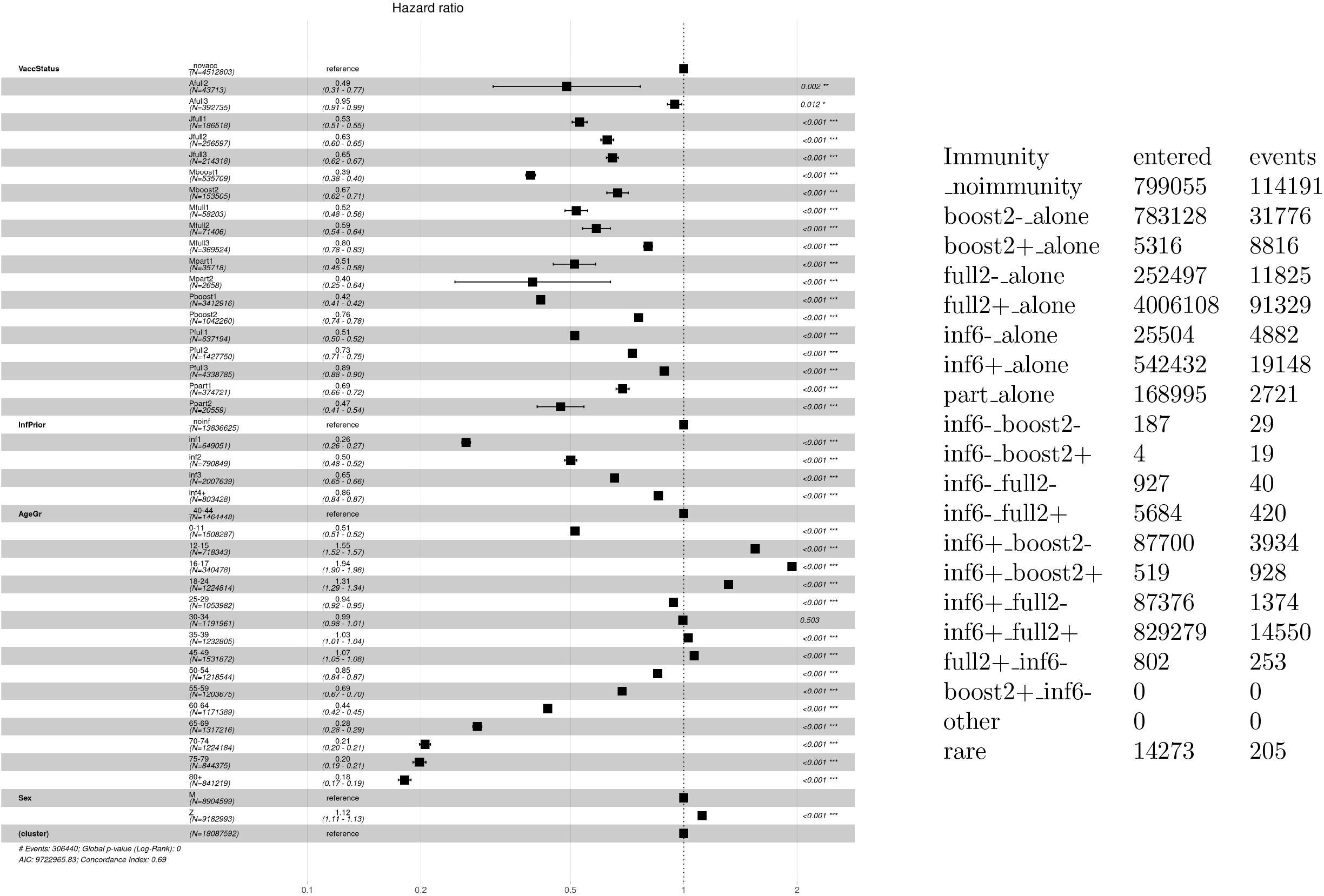

### 12 Infections by Vaccines - Delta

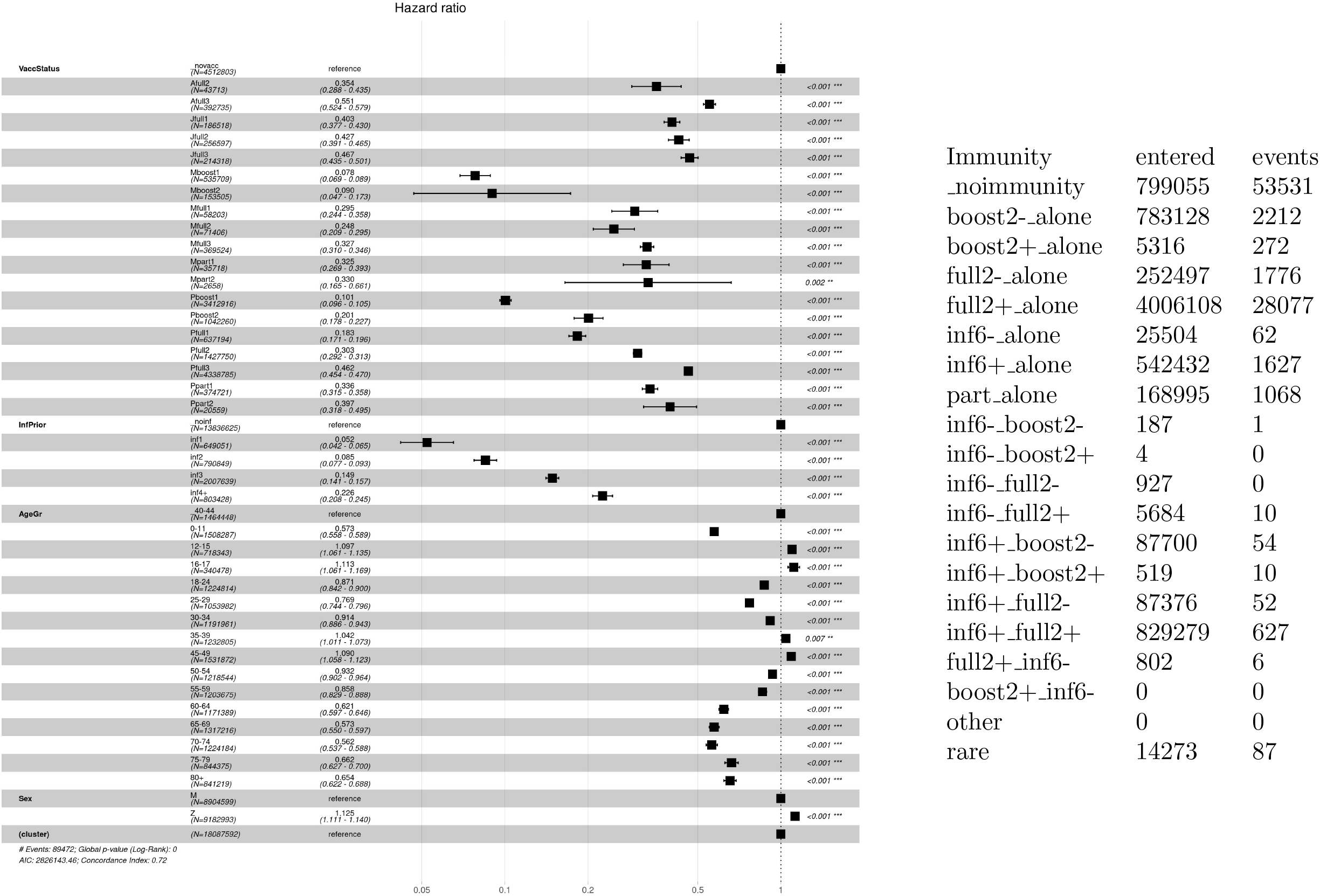

### 13 Hospitalizations by Vaccines - Omicron

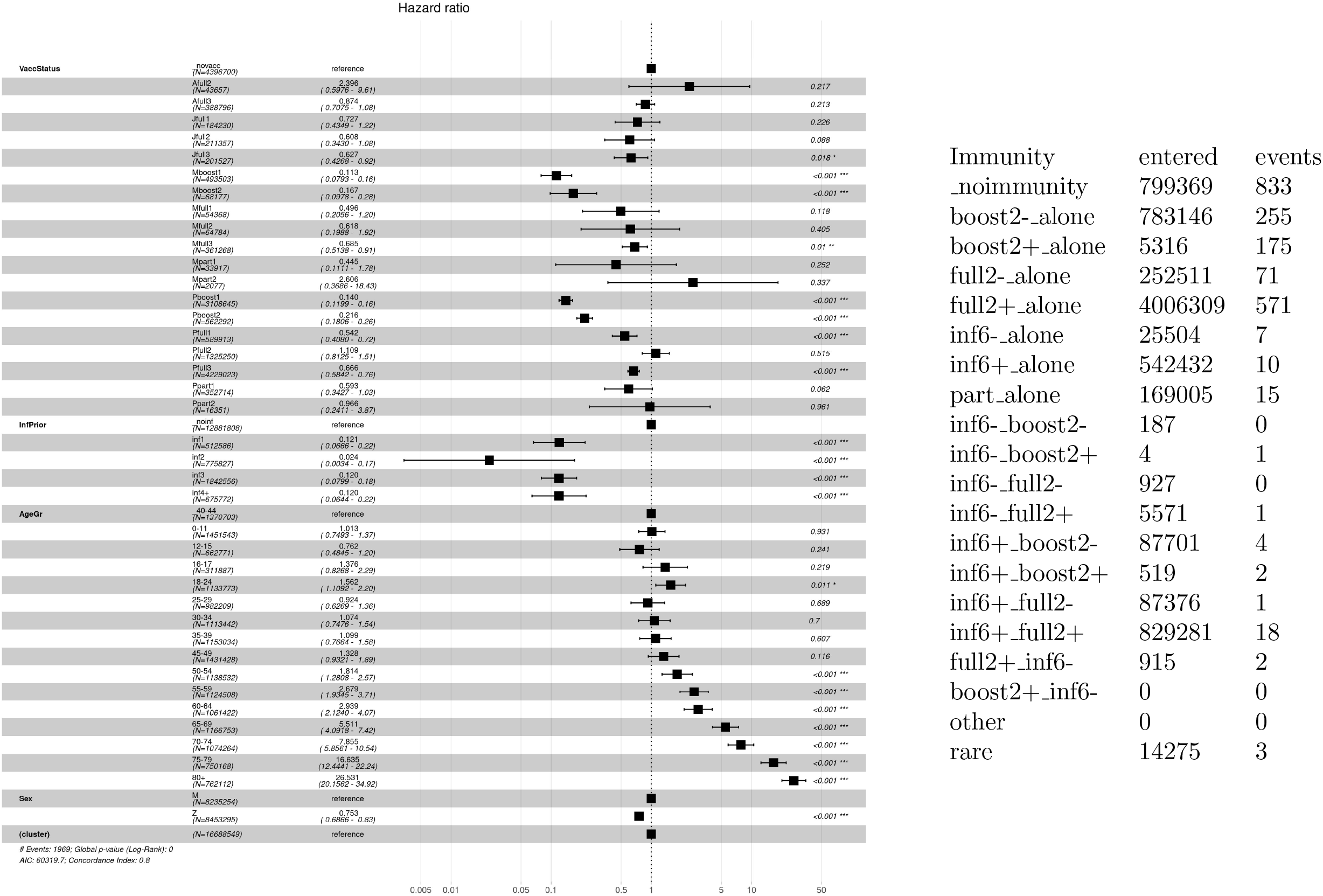

### 14 Hospitalizations by Vaccines - Delta

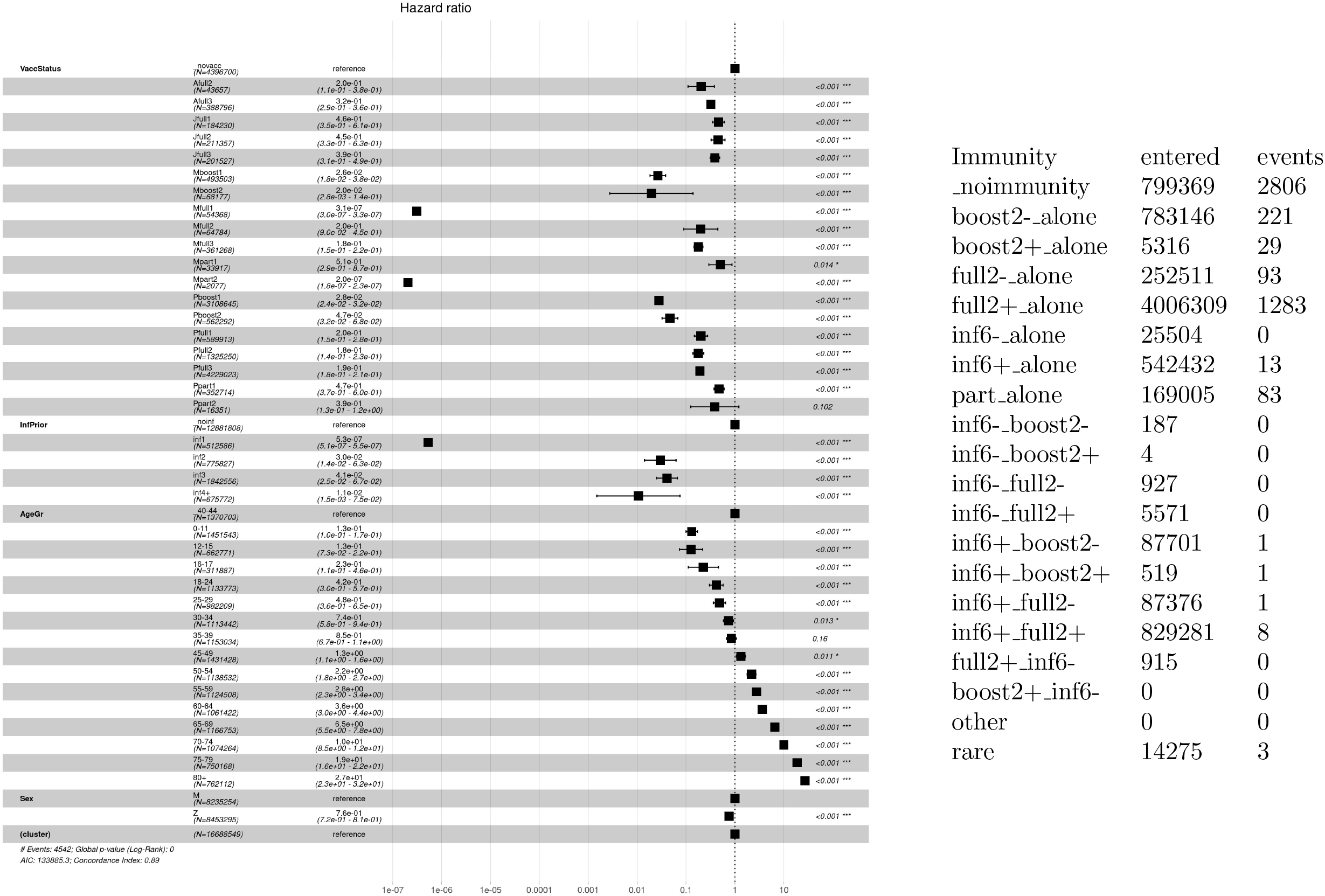

### 15 Hospitalizaton given Infection

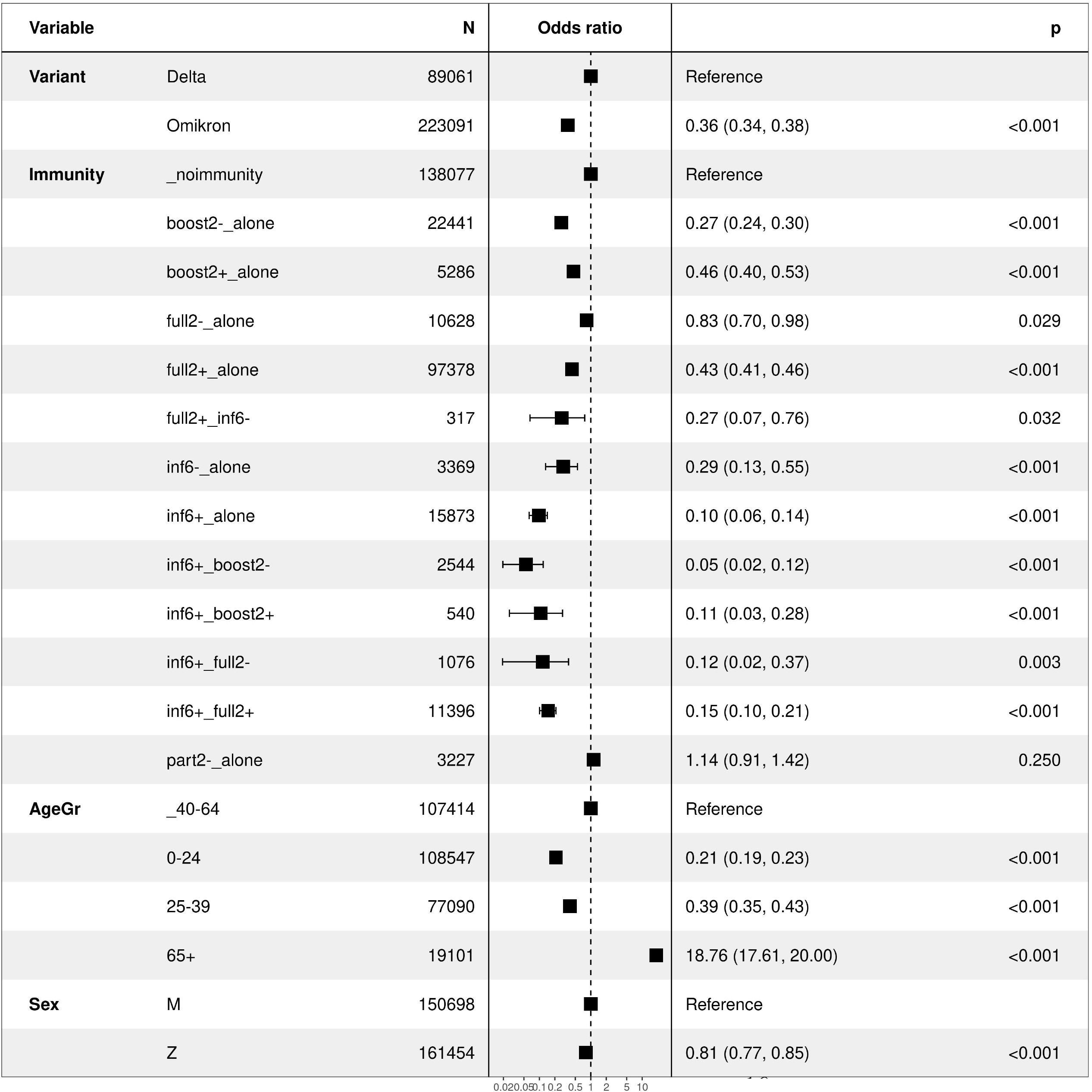

| Variable |  | N | Odds ratio | p |  |
| --- | --- | --- | --- | --- | --- |
| Variant | Delta | 89061 |  | Reference |  |
|  | Omikron | 223091 |  | 0.36 (0.34, 0.38) | <0.001 |
| Immunity | _noimmunity | 138077 |  | Reference |  |
|  | boost2-_alone | 22441 |  | 0.27 (0.24, 0.30) | <0.001 |
|  | boost2+_alone | 5286 |  | 0.46 (0.40, 0.53) | <0.001 |
|  | full2-_alone | 10628 |  | 0.83 (0.70, 0.98) | 0.029 |
|  | full2+_alone | 97378 |  | 0.43 (0.41, 0.46) | <0.001 |
|  | full2+_inf6- | 317 |  | 0.27 (0.07, 0.76) | 0.032 |
|  | inf6-_alone | 3369 |  | 0.29 (0.13, 0.55) | <0.001 |
|  | inf6+_alone | 15873 |  | 0.10 (0.06, 0.14) | <0.001 |
|  | inf6+_boost2- | 2544 |  | 0.05 (0.02, 0.12) | <0.001 |
|  | inf6+_boost2+ | 540 |  | 0.11 (0.03, 0.28) | <0.001 |
|  | inf6+_full2- | 1076 |  | 0.12 (0.02, 0.37) | 0.003 |
|  | inf6+_full2+ | 11396 |  | 0.15 (0.10, 0.21) | <0.001 |
|  | part2-_alone | 3227 |  | 1.14 (0.91, 1.42) | 0.250 |
| AgeGr | _40-64 | 107414 |  | Reference |  |
|  | 0-24 | 108547 |  | 0.21 (0.19, 0.23) | <0.001 |
|  | 25-39 | 77090 |  | 0.39 (0.35, 0.43) | <0.001 |
|  | 65+ | 19101 |  | 18.76 (17.61, 20.00) | <0.001 |
| Sex | M | 150698 |  | Reference |  |
|  | Z | 161454 |  | 0.81 (0.77, 0.85) | <0.001 |

### 16 Oxygen given Infection

| Variable |  | N | Odds ratio | p |  |
| --- | --- | --- | --- | --- | --- |
| Variant | Delta | 89038 | ■ | Reference |  |
|  | Omikron | 223041 | ■ | 0.24 (0.22, 0.26) | <0.001 |
| Immunity | _noimmunity | 138077 | ■ | Reference |  |
|  | boost2-_alone | 22441 | ■ | 0.21 (0.19, 0.25) | <0.001 |
|  | boost2+_alone | 5286 | ■ | 0.37 (0.29, 0.45) | <0.001 |
|  | full2-_alone | 10628 | ■ | 0.54 (0.41, 0.70) | <0.001 |
|  | full2+_alone | 97378 | ■ | 0.35 (0.32, 0.38) | <0.001 |
|  | full2+_inf6- | 314 | ■ | 0.00 (0.00, 0.00) | 1.0 |
|  | inf6-_alone | 3361 | ■ | 0.00 (0.00, 0.00) | 1.0 |
|  | inf6+_alone | 15850 | ■ | 0.01 (0.00, 0.04) | <0.001 |
|  | inf6+_boost2- | 2539 | ■ | 0.00 (0.00, 0.00) | 1.0 |
|  | inf6+_boost2+ | 537 | ■ | 0.00 (0.00, 0.00) | 1.0 |
|  | inf6+_full2- | 1074 | ■ | 0.00 (0.00, 0.00) | 1.0 |
|  | inf6+_full2+ | 11367 | ■ | 0.00 (0.00, 0.00) | 0.9 |
|  | part2-_alone | 3227 | ■ | 0.78 (0.57, 1.06) | 0.1 |
| AgeGr | _40-64 | 107391 | ■ | Reference |  |
|  | 0-24 | 108545 | ■ | 0.04 (0.03, 0.06) | <0.001 |
|  | 25-39 | 77082 | ■ | 0.20 (0.17, 0.24) | <0.001 |
|  | 65+ | 19061 | ■ | 18.44 (17.01, 20.02) | <0.001 |
| Sex | M | 150658 | ■ | Reference |  |
|  | Z | 161421 | ■ | 0.72 (0.67, 0.77) | <0.001 |

1e-2861e-2291e-1721e-1151e-58 0.1

### 17 ICU given Infection

| Variable |  | N | Odds ratio | p |  |
| --- | --- | --- | --- | --- | --- |
| Variant | Delta | 89061 | ■ | Reference |  |
|  | Omikron | 223091 | ■ | 0.24 (0.21, 0.28) | <0.001 |
| Immunity | _noimmunity | 138077 | ■ | Reference |  |
|  | boost2-_alone | 22441 | ■ | 0.30 (0.23, 0.38) | <0.001 |
|  | boost2+_alone | 5286 | ■ | 0.65 (0.46, 0.90) | 0.012 |
|  | full2-_alone | 10628 | ■ | 0.43 (0.25, 0.70) | 0.001 |
|  | full2+_alone | 97378 | ■ | 0.33 (0.28, 0.38) | <0.001 |
|  | full2+_inf6- | 317 | ■ | 0.00 (0.00, 0.00) | 0.979 |
|  | inf6-_alone | 3369 | ■ | 0.32 (0.02, 1.45) | 0.262 |
|  | inf6+_alone | 15873 | ■ | 0.15 (0.05, 0.32) | <0.001 |
|  | inf6+_boost2- | 2544 | ■ | 0.00 (0.00, 0.00) | 0.943 |
|  | inf6+_boost2+ | 540 | ■ | 0.00 (0.00, 0.00) | 0.973 |
|  | inf6+_full2- | 1076 | ■ | 0.00 (0.00, 0.00) | 0.965 |
|  | inf6+_full2+ | 11396 | ■ | 0.04 (0.00, 0.16) | <0.001 |
|  | part2-_alone | 3227 | ■ | 0.78 (0.44, 1.28) | 0.359 |
| AgeGr | _40-64 | 107414 | ■ | Reference |  |
|  | 0-24 | 108547 | ■ | 0.09 (0.06, 0.12) | <0.001 |
|  | 25-39 | 77090 | ■ | 0.19 (0.14, 0.26) | <0.001 |
|  | 65+ | 19101 | ■ | 10.24 (8.94, 11.76) | <0.001 |
| Sex | M | 150698 | ■ | Reference |  |
|  | Z | 161454 | ■ | 0.55 (0.48, 0.62) | <0.001 |

1e-22 11e-17 71e-13 31e-8 9 1e-4 5 0.1

### 18 Oxagen given Hospitalization

18 Oxygen given Hospitalization
| Variable |  | N | Odds ratio | p |  |
| --- | --- | --- | --- | --- | --- |
| Variant | Delta | 4521 | ■ | Reference |  |
|  | Omikron | 2235 | ■ | 0.44 (0.39, 0.50) | <0.001 |
| Immunity | _noimmunity | 3788 | ■ | Reference |  |
|  | boost2-_alone | 516 | ■ | 0.47 (0.38, 0.57) | <0.001 |
|  | boost2+_alone | 246 | ■ | 0.45 (0.34, 0.60) | <0.001 |
|  | full2-_alone | 175 | ■ | 0.39 (0.28, 0.55) | <0.001 |
|  | full2+_alone | 1920 | ■ | 0.56 (0.50, 0.63) | <0.001 |
|  | inf6+_alone | 1 | ■ | 147714.80 (0.00, NA) | 0.952 |
|  | inf6+_full2+ | 1 | ■ | 0.00 (NA, 10308582286697.01) | 0.959 |
|  | part2-_alone | 109 | ■ | 0.54 (0.36, 0.82) | 0.003 |
| AgeGr | _40-64 | 1568 | ■ | Reference |  |
|  | 0-24 | 445 | ■ | 0.12 (0.09, 0.16) | <0.001 |
|  | 25-39 | 484 | ■ | 0.32 (0.25, 0.40) | <0.001 |
|  | 65+ | 4259 | ■ | 1.64 (1.45, 1.86) | <0.001 |
| Sex | M | 3330 | ■ | Reference |  |
|  | Z | 3426 | ■ | 0.79 (0.71, 0.88) | <0.001 |

1e-12 1e-07 0.01 1000 1e+08 1e+13

### 19 ICU given Hospitalization

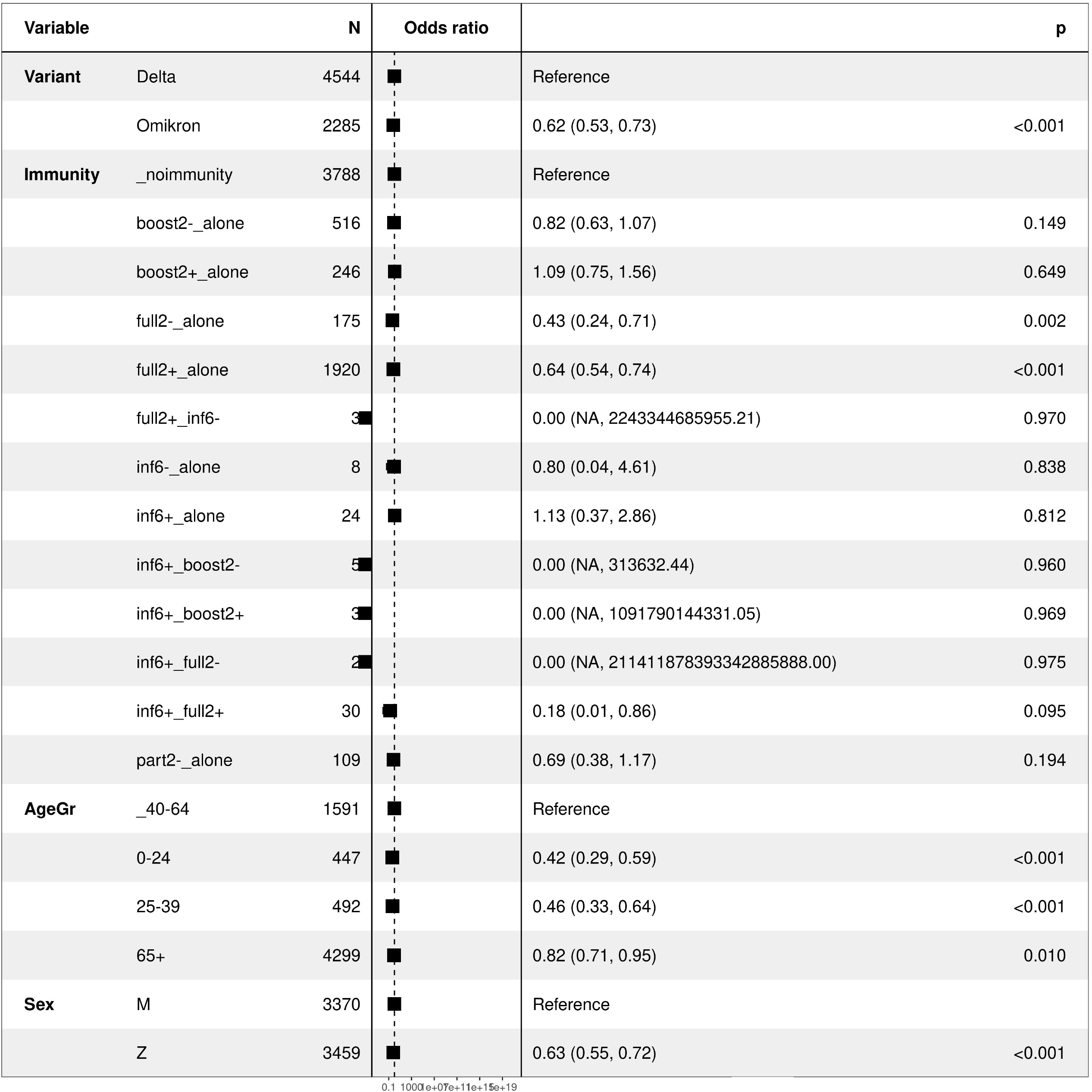

